# COVID-19 symptoms at hospital admission vary with age and sex: ISARIC multinational study

**DOI:** 10.1101/2020.10.26.20219519

**Authors:** Mark G Pritchard, ISARIC Clinical Characterisation Group

**Affiliations:** Centre for Tropical Medicine and Global Health, Nuffield Department of Medicine, University of Oxford, Oxford, UK

**Keywords:** age group, COVID-19, SARS-CoV-2, sex, symptoms, diagnosis, case definition

## Abstract

**Background:** The ISARIC prospective multinational observational study is the largest cohort of hospitalized patients with COVID-19. We present relationships of age, sex, and nationality to presenting symptoms.

**Methods:** International, prospective observational study of 60⍰109 hospitalized symptomatic patients with laboratory-confirmed COVID-19 recruited from 43 countries between 30 January and 3 August 2020. Logistic regression was performed to evaluate relationships of age and sex to published COVID-19 case definitions and the most commonly reported symptoms.

**Results:** ‘Typical’ symptoms of fever (69%), cough (68%) and shortness of breath (66%) were the most commonly reported. 92% of patients experienced at least one of these. Prevalence of typical symptoms was greatest in 30-to 60-year-olds (respectively 80%, 79%, 69%; at least one 95%). They were reported less frequently in children (≤18 years: 69%, 48%, 23%; 85%), older adults (≥70 years: 61%, 62%, 65%; 90%), and women (66%, 66%, 64%; 90%; vs men 71%, 70%, 67%; 93%). The most common atypical presentation under 60 years of age was nausea and vomiting, and over 60 years was confusion. Regression models showed significant differences in symptoms with sex, age and country.

**Interpretation:** Adults over 60 and children admitted to hospital with COVID-19 are less likely to present with typical symptoms. Nausea and vomiting are common atypical presentations under 30 years. Confusion is a frequent atypical presentation of COVID-19 in adults over 60 years. Women are less likely to experience typical symptoms than men.

**Summary:** Adults over 60 and children admitted to hospital with COVID-19 are less likely to have typical symptoms. Nausea and vomiting are common atypical presentations under 30 and confusion over 60. Women are less likely to experience typical symptoms than men.

## Background

Despite the pandemic’s immense human cost, enormous economic toll, and extensive research response, the precise clinical characteristics of COVID-19 remain unclear [1]. At the start of the outbreak, COVID-19 was broadly characterised as a severe respiratory illness presenting with fever, cough and an atypical pneumonia [2-5]. Altered sense of taste and smell have since been found to be strongly associated with the disease [6, 7]. However, a review of 77 observational studies found substantial proportions of patients presenting with less typical symptoms [8].

The World Health Organization’s (WHO) clinical criteria for suspected COVID-19 is either a combination of acute fever and cough, or a combination of three or more of fever, cough, general weakness and fatigue, headache, myalgia, sore throat, coryza, dyspnoea, anorexia, nausea and vomiting, diarrhoea, and altered mental status [9]. Clinical criteria from the Centers for Disease Control and Prevention in the United States are at least two of fever, chills, rigors, myalgia, headache, sore throat, and new olfactory and taste disorder, or at least one of cough, shortness of breath or difficulty breathing [10]. Public Health England’s definition of possible COVID-19 is individuals with a new cough or temperature ≥37.8 °C, or a loss or change in sense of smell or taste [11]. The European Centre for Disease Prevention and Control clinical criteria are at least one of cough, fever, shortness of breath, or sudden onset anosmia, ageusia or dysgeusia [12].

Defining presenting symptoms of COVID-19 is further complicated by clinical experience suggesting that patients frequently present with atypical symptoms other than cough, fever and shortness of breath. This variation in the clinical characterisation of COVID-19 is problematic, as case definitions are used to guide clinical diagnosis, disease surveillance, and public health interventions.

The International Severe Acute Respiratory and Emerging Infection Consortium (ISARIC)/WHO Clinical Characterisation Protocol for Severe Emerging Infections is a standardized protocol for investigation of severe acute infections by pathogens of public health interest [13, 14]. Here we present an analysis of how symptoms of patients admitted to hospital with confirmed COVID-19 vary by age and sex.

## Materials and methods

### Study population, variables and measurement

Patients of any age admitted to hospital with suspected or confirmed COVID-19 were eligible for recruitment. This analysis was limited to patients admitted to hospital between 30 January and 3 August 2020 with symptomatic COVID-19, confirmed according to sites’ local laboratory methods. We excluded asymptomatic patients admitted to hospital solely for isolation, and patients admitted for other conditions who subsequently developed COVID-19 symptoms. Variables used in this analysis were age, sex at birth, symptoms, date of symptoms onset, SARS-CoV-2 confirmation, and country of recruitment. To allow proportions to be calculated with a reliable denominator, only symptoms specified on the case report forms were included in this analysis.

This observational study required no change to clinical management and permitted enrolment in other research. The study was approved by the World Health Organization Ethics Review Committee (RPC571 and RPC572). Local ethics approval was obtained for each participating country and site according to local requirements. Informed consent was obtained where required by local ethics committees. In many jurisdictions, ethics committees and public health authorities approved a waiver of consent.

### Study design and setting

The ISARIC cohort is an international prospective observational study of patients admitted to hospital with COVID-19. Data were collected via electronic ‘Core’ and ‘Rapid’ ISARIC case report forms [15], and through aligned forms by ISARIC-4C Coronavirus Clinical Characterisation Consortium in the United Kingdom [16] and the COVID-19 Critical Care Consortium [17]. Investigators from 41 countries used Research Electronic Data Capture (REDCap, version 8.11.11, Vanderbilt University, Nashville, Tenn.) to contribute their data to a central database hosted by the University of Oxford. Additional data were submitted by investigators not using the University of Oxford REDCap database from Malaysia, Russia [18], and Spain. The primary objective of this analysis was to investigate how symptoms of patients admitted to hospital with confirmed COVID-19 vary by age and sex. Secondarily we investigated how sensitivity of clinical case definitions varied among these populations, and explored heterogeneity among countries.

### Analysis

Data were converted to Study Data Tabulation Model (version 1.7, Clinical Data Interchange Standards Consortium, Austin, Tex.). We excluded patients who had all symptoms recorded as missing or unknown, and those with missing age, sex, country or onset date. Continuous variables were expressed as median with interquartile range (IQR), and categorical variables as counts with percentages. We tested for differences between female and male patients using Wilcoxon rank-sum tests for continuous variables and chi-square tests for categorical variables. We grouped patients into ten-year age bands (with a single group ≥90 years). We plotted most frequently reported symptoms by age group, presenting missing data as a third category. We collated symptoms according to four sets of clinical criteria, based on published criteria [9-12] modified to omit symptoms with large numbers of missing data:

1. Fever plus cough; or any three of fever, cough, fatigue, headache, myalgia, sore throat, rhinorrhoea, shortness of breath, nausea and vomiting, diarrhoea, and confusion;
2. Cough or shortness of breath; or any two of fever, myalgia, headache, and sore throat;
3. Cough or fever;
4. At least one of cough, fever, and shortness of breath.

Patients with missing details of cough, fever or shortness of breath were omitted from the composite groups; patients missing details of symptoms included in the lists of criteria 1 and 2 were classified according to their non-missing symptoms. We plotted proportions of patients meeting each set of criteria by ten-year age group with 95% confidence intervals (CI) calculated using the Clopper–Pearson method.

We used logistic regression to identify associations of age and sex with the twelve most prevalent symptoms. Age group and sex were included as fixed effects, with country as a random intercept. To display heterogeneity between countries on the same scale as the fixed effects, we plotted median odds ratios (MOR), which quantify variation between countries by comparing odds of an outcome between randomly chosen persons in different clusters who share covariates [19]. MORs are defined as a comparison of the greater propensity group to the lower propensity group, so lie in the range one to infinity [19].

79% of patients were recruited in a single country. As a sensitivity analysis, we repeated the analysis excluding patients from this country. Finally, we plotted age-stratified symptom frequencies for each country with at least 250 patients.

No minimum sample size was calculated. All significance tests were two-tailed. Analyses were performed using R (version 4.0.3, R Foundation for Statistical Computing, Vienna, Austria) with packages including *binom, Epi, ggplot2, lme4, sjstats, tableone*, and *tidyverse*.

## Results

Data were available for 99⍰623 patients. We excluded 24⍰336 who did not have documented SARS-CoV-2 confirmation, 3290 with missing data, and 5794 who developed COVID-19 after admission to hospital. 6094 patients were admitted to hospital with asymptomatic COVID-19, with the greatest proportion in the age-band 10 to 20 years (46%). We included 60⍰109 patients (Figure 1), recruited from 394 sites in 43 countries (Supplementary Table 1), in this analysis. The median age of included patients was 70 years (IQR 54–82; Table 1). 929 (1.5%) were 18 years old or younger. Age distribution of patients varied among countries, between a median of 10 years in Poland, and 73 years in the United Kingdom (Supplementary Figure 1). 34⍰641 (58%) patients were male.

**Table 1.**
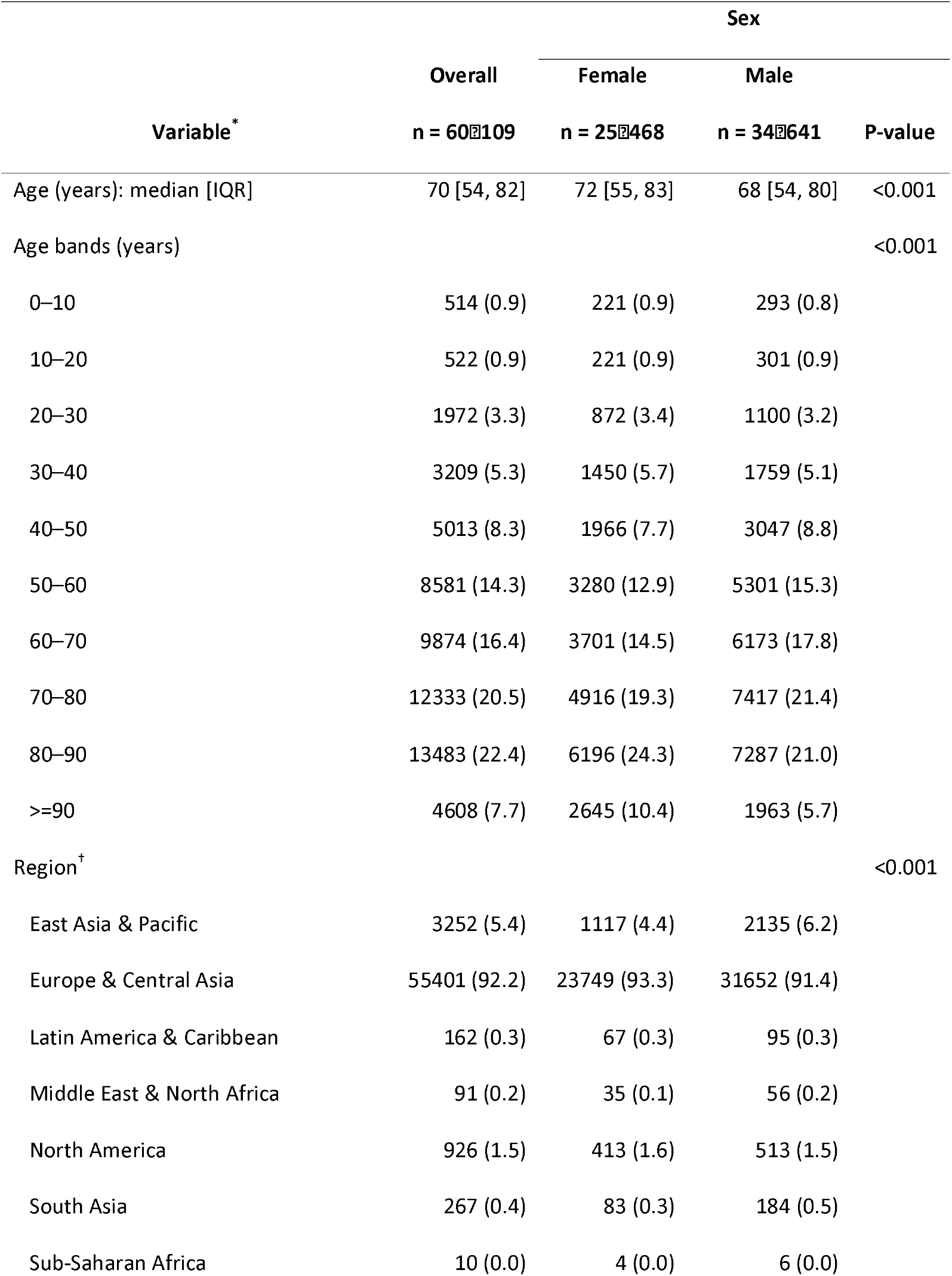
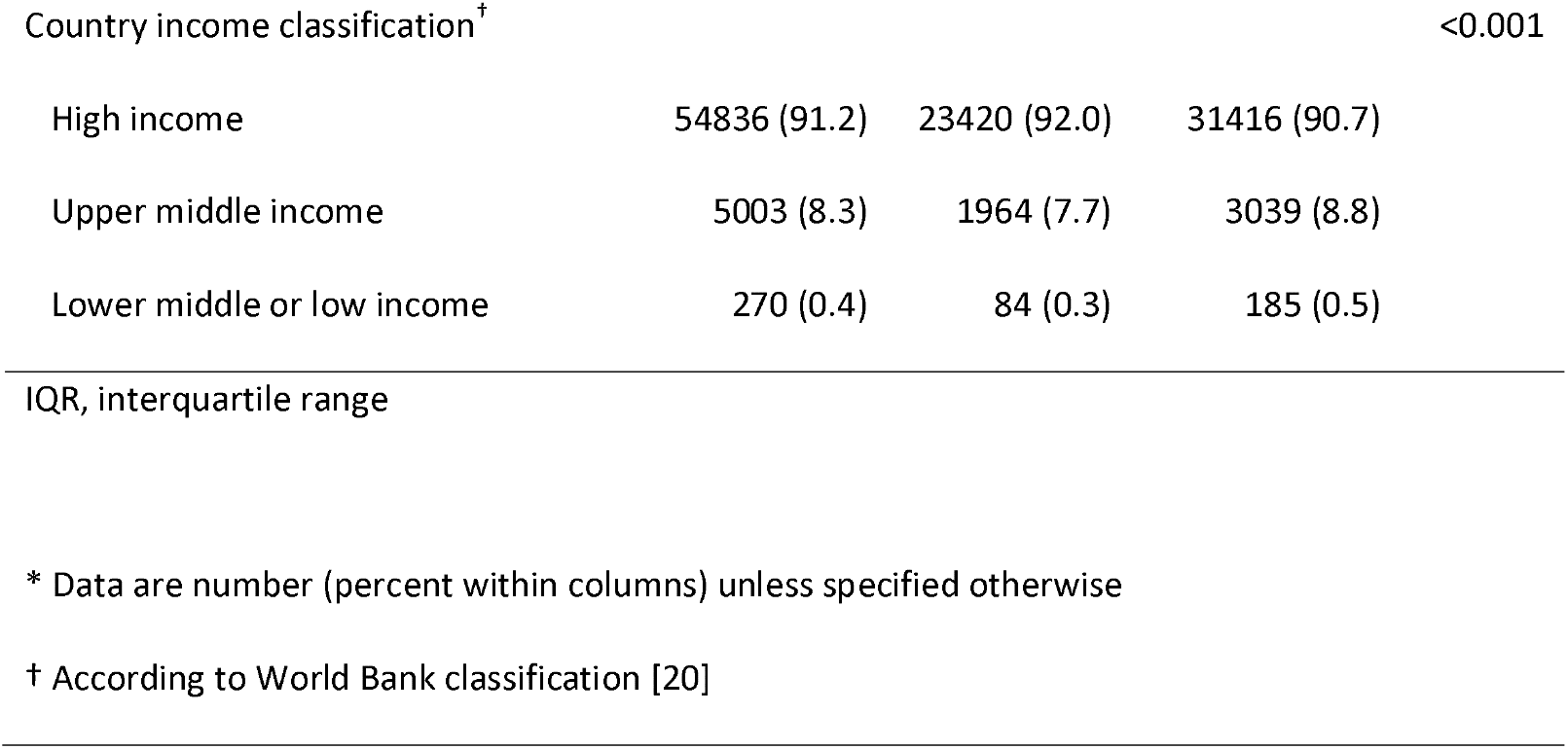
Patient demographics.

**Figure 1.**
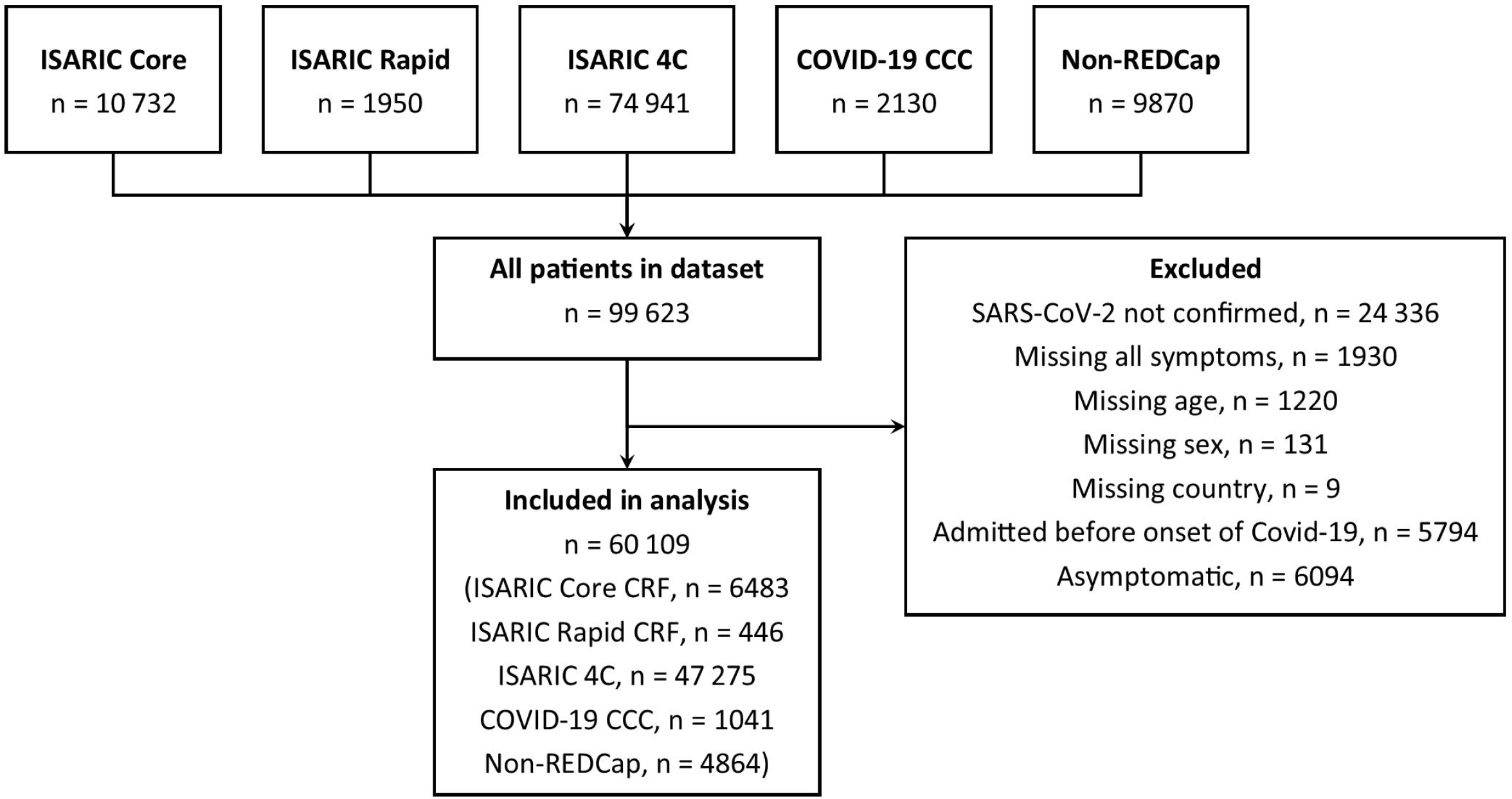
Flow of participants in this analysis. 4C, Coronavirus Clinical Characterisation Consortium; CCC, Critical Care Consortium; ISARIC, International Severe Acute Respiratory and emerging Infection Consortium; REDCap, Research Electronic Data Capture; SARS-CoV-2, severe acute respiratory syndrome coronavirus-2.

The most frequently reported symptoms were fever, cough and shortness of breath (Table 2). These symptoms were each more prevalent in male patients, whereas less typical symptoms such as confusion, nausea and vomiting, diarrhoea, chest pain, headache and abdominal pain were more prevalent in female patients. The greatest sex-related difference was for nausea and vomiting, reported by 23% of female patients but only 16% of male patients. For most symptoms, the greatest prevalence was reported in adults aged between 30 and 60 years, decreasing toward extremities of age (Figure 2). Frequency of confusion increased with age. Large numbers of patients had missing data for anorexia, severe dehydration, altered sense of taste and smell, and inability to walk as these were not included on all case report forms. Altered sense of taste and smell, which we had excluded from the composite criteria, were experienced by only 7.4% and 6.2% respectively of patients with non-missing data.

**Table 2.**
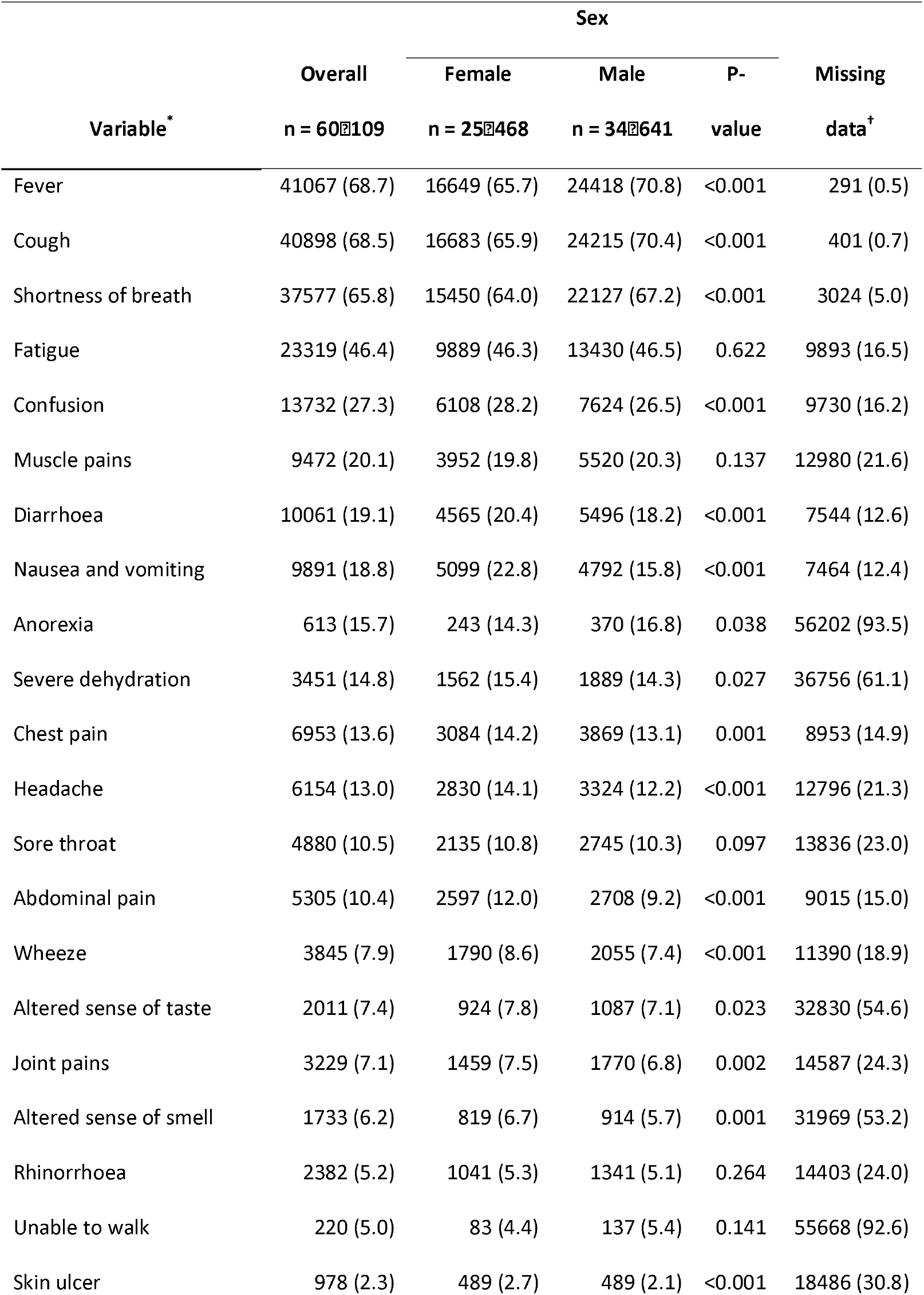
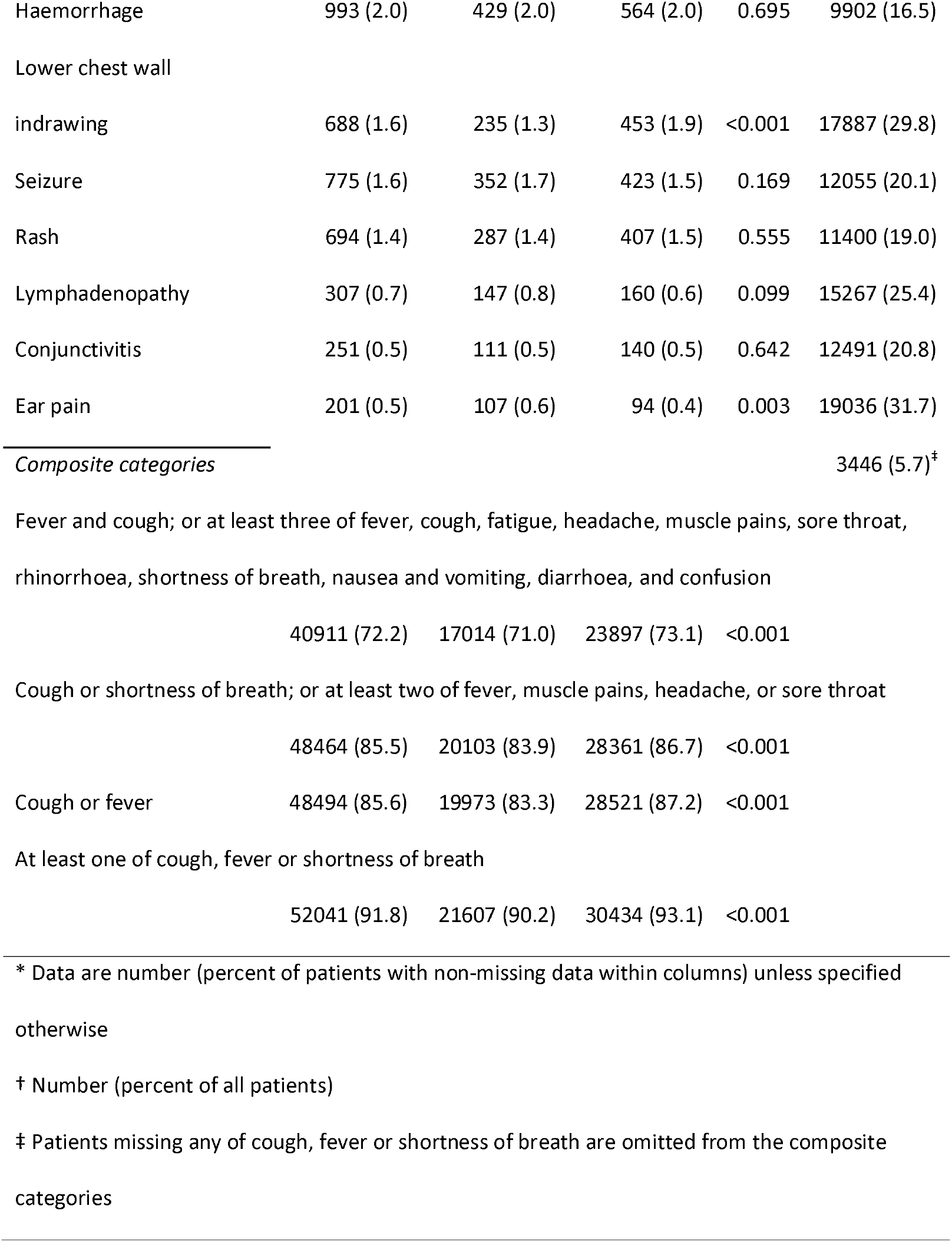
Symptoms at presentation to hospital with COVID-19.

**Figure 2.**
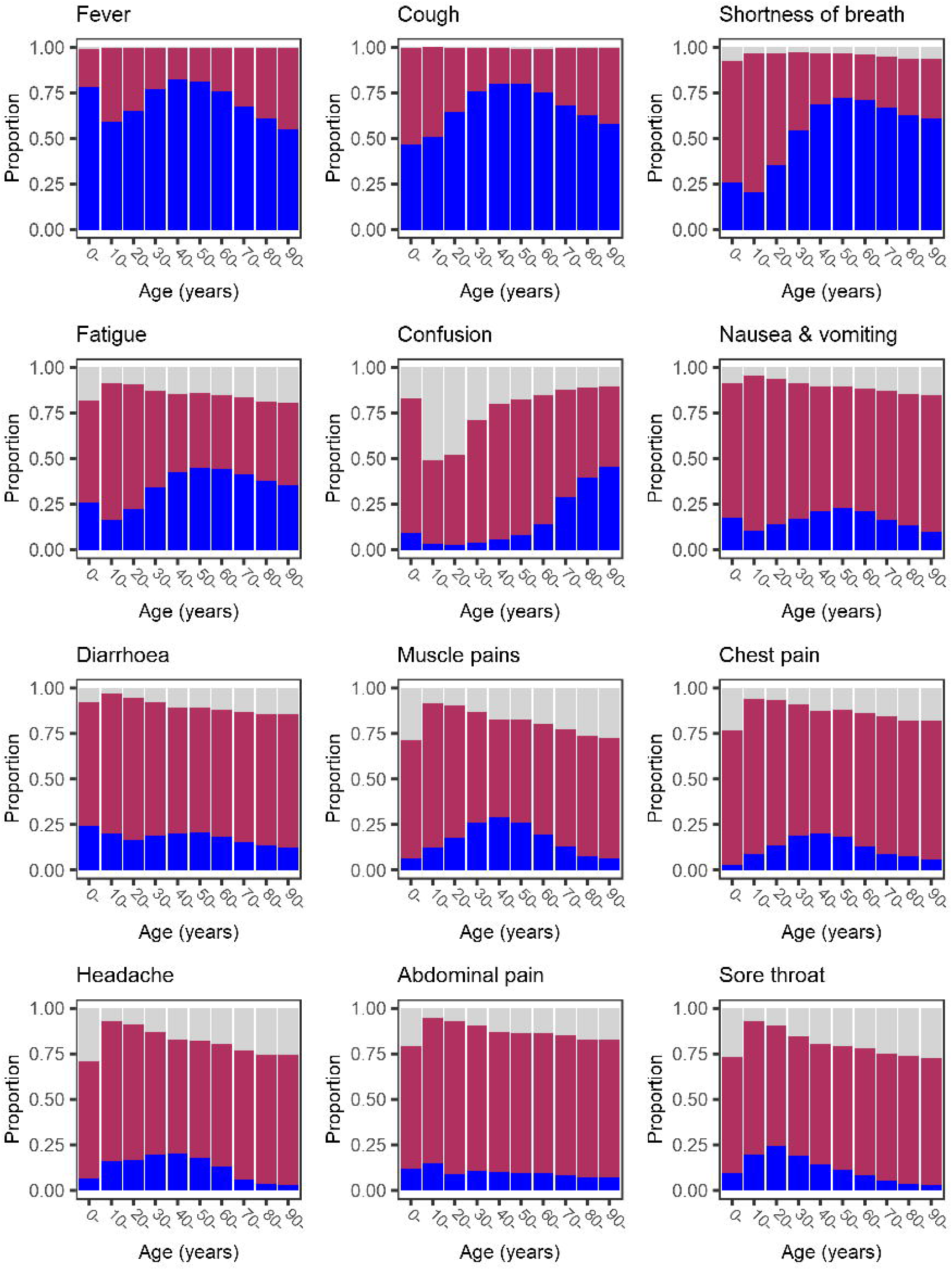
Age-specific prevalence of symptoms at hospital admission. Dark blue bars show symptom present, maroon bars show symptom absent, pale grey bars show missing data.

Data on cough, fever or shortness of breath were missing for 3446 patients. The composite clinical criteria were calculated for the remaining 56⍰663 patients. Each set of criteria was met by a greater proportion of patients aged 30 to 60 years than those toward either extreme of age (Figure 3). The criteria based on WHO’s clinical criteria [9] (fever plus cough; or any three of fever, cough, fatigue, headache, myalgia, sore throat, rhinorrhoea, shortness of breath, nausea and vomiting, diarrhoea, and confusion) were met by 40⍰911 (72%) patients, but only 51% of those aged 18 years and under, and 67% of those aged 70 years or over. The most sensitive criteria were at least one of cough, fever and shortness of breath, met by 52⍰041 (92%) participants. These criteria were met by 85% aged 18 years and under, and 90% of those aged 70 years or over. Each set of criteria were met by a greater proportion of male than female patients (Table 2).

**Figure 3.**
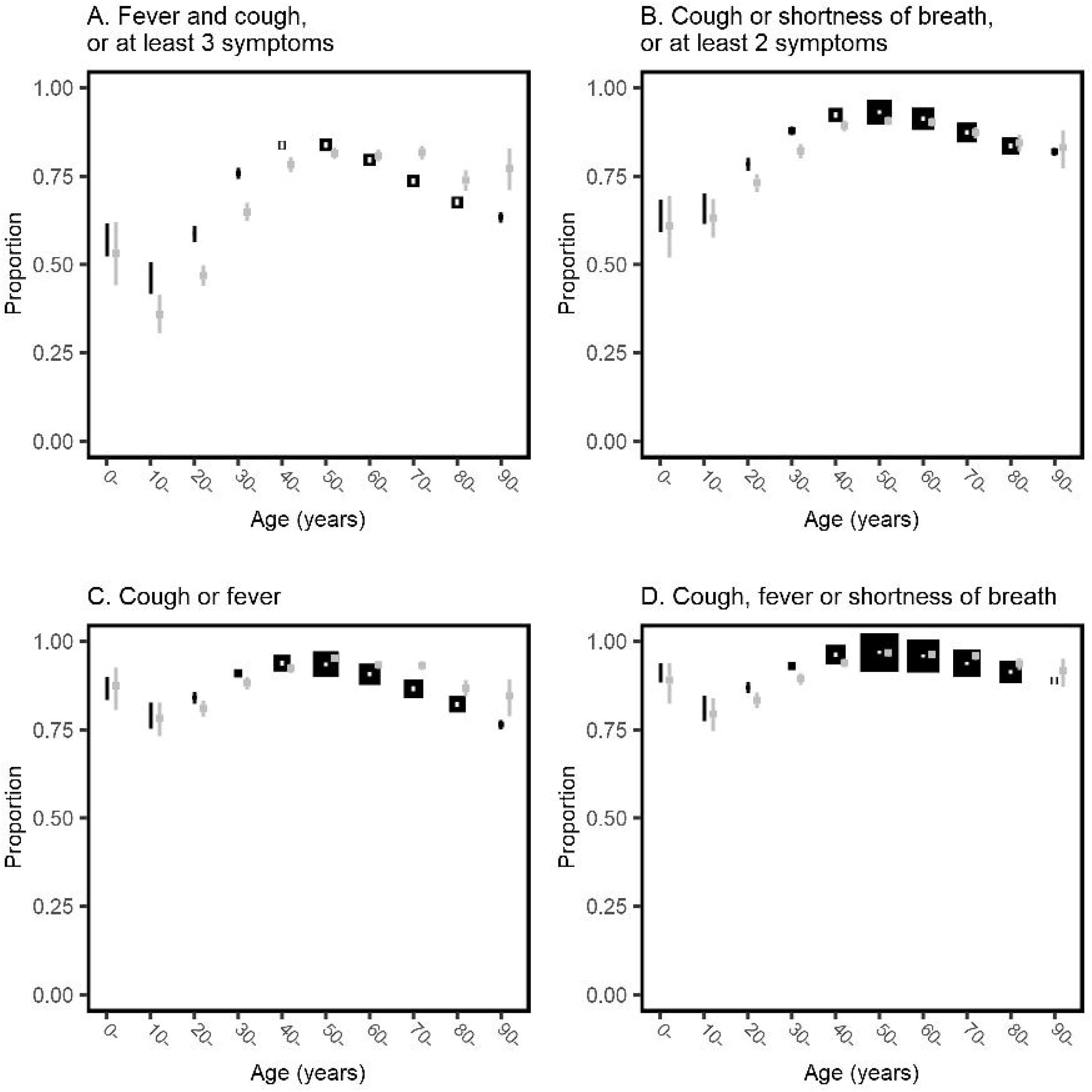
Proportions meeting clinical criteria at hospital admission stratified by 10-year age band. Black boxes show the proportion of individuals, with error bars showing 95% confidence intervals calculated using the Clopper–Pearson method. The size of each box is inversely proportional to the variance, so larger boxes indicate greater certainty. Grey boxes with 95% confidence intervals show the proportions in the sensitivity analysis excluding patients recruited in the United Kingdom. In panel A, the three symptoms are from the list of fever, cough, fatigue, headache, myalgia, sore throat, rhinorrhoea, shortness of breath, nausea and vomiting, diarrhoea, and confusion; in panel B the two symptoms are from the list fever, myalgia, headache, and sore throat. Patients with missing data for cough, fever or shortness of breath are excluded from all four plots.

For the 4622 patients whose symptoms did not meet any assessed case definitions, the most frequent symptom was confusion (47%; Table 3). This increased with age to 66% of those aged 90 years or older. Nausea and vomiting, and abdominal pain were the most common symptoms for people less than 60 years old who had not met any of the case definitions.

**Table 3.** Symptoms reported for patients meeting none of the clinical case definitions.

| Symptom * | Age group (years) |  |  |  |  |  |  |  |  |  |  | Missing data |
| --- | --- | --- | --- | --- | --- | --- | --- | --- | --- | --- | --- | --- |
|  | Overall<br>n = 4622 | 0–<br>n = 44 | 10–<br>n = 100 | 20–<br>n = 269 | 30–<br>n = 256 | 40–<br>n = 217 | 50–<br>n = 309 | 60–<br>n = 484 | 70–<br>n = 922 | 80–<br>n = 1406 | 90–<br>n = 615 |  |
| Confusion | 1917 (46.9) | 4 (9.8) | 2 (4.2) | 7 (6.2) | 9 (5.8) | 23 (14.1) | 57 (21.3) | 155 (35.1) | 474 (52.8) | 785 (57.7) | 401 (66.4) | 531 (11.5) |
| Fatigue | 1245 (29.1) | 9 (23.1) | 10 (10.3) | 30 (11.3) | 37 (15.2) | 42 (20.7) | 83 (28.1) | 148 (33.2) | 284 (33.3) | 444 (34.4) | 158 (28.6) | 340 (7.4) |
| Severe dehydration | 472 (22.6) | 1 (5.0) | 1 (4.3) | 4 (8.3) | 2 (2.9) | 7 (11.3) | 18 (13.1) | 40 (18.0) | 110 (24.8) | 201 (27.1) | 88 (27.6) | 2535 (54.8) |
| Nausea and vomiting | 995 (22.2) | 19 (44.2) | 24 (24.0) | 44 (16.4) | 59 (23.4) | 77 (36.0) | 94 (31.2) | 127 (27.1) | 210 (23.4) | 245 (18.3) | 96 (16.2) | 145 (3.1) |
| Abdominal pain | 812 (18.5) | 9 (22.5) | 22 (22.4) | 52 (19.5) | 67 (26.6) | 54 (25.8) | 80 (27.0) | 107 (23.1) | 173 (19.6) | 184 (13.9) | 64 (11.2) | 223 (4.8) |
| Diarrhoea | 612 (13.8) | 8 (19.0) | 6 (6.1) | 26 (9.7) | 25 (10.0) | 31 (14.8) | 46 (15.5) | 94 (20.3) | 136 (15.2) | 177 (13.4) | 63 (10.9) | 201 (4.3) |
| Headache | 347 (8.3) | 2 (6.1) | 13 (13.4) | 48 (18.4) | 39 (16.0) | 47 (23.2) | 32 (11.0) | 45 (10.3) | 52 (6.3) | 47 (3.8) | 22 (4.1) | 454 (9.8) |
| Muscle pains | 301 (7.3) | 4 (11.8) | 5 (5.2) | 19 (7.2) | 28 (11.5) | 24 (12.1) | 27 (9.4) | 39 (9.0) | 54 (6.6) | 65 (5.3) | 36 (6.8) | 482 (10.4) |
| Joint pains | 293 (7.1) | 3 (9.1) | 4 (4.2) | 9 (3.4) | 8 (3.3) | 13 (6.5) | 14 (4.9) | 27 (6.2) | 49 (6.0) | 109 (8.8) | 57 (10.7) | 473 (10.2) |
| Chest pain | 299 (6.8) | 0 (0.0) | 1 (1.0) | 12 (4.5) | 15 (6.0) | 21 (9.8) | 44 (14.6) | 49 (10.6) | 51 (5.8) | 79 (6.0) | 27 (4.7) | 209 (4.5) |
| Sore throat | 277 (6.7) | 4 (11.1) | 24 (25.0) | 83 (31.7) | 62 (25.1) | 39 (19.3) | 16 (5.5) | 18 (4.1) | 17 (2.1) | 9 (0.7) | 5 (1.0) | 460 (10.0) |
| Haemorrhage | 233 (5.5) | 0 (0.0) | 1 (1.1) | 5 (2.1) | 10 (4.3) | 12 (6.0) | 14 (4.8) | 22 (4.9) | 45 (5.2) | 84 (6.5) | 40 (7.0) | 347 (7.5) |
| Altered sense of smell | 149 (4.9) | 1 (4.0) | 20 (24.1) | 48 (22.7) | 30 (16.0) | 17 (11.4) | 11 (5.1) | 9 (2.8) | 4 (0.7) | 9 (1.0) | 0 (0.0) | 1584 (34.3) |
| Rhinorrhoea | 188 (4.5) | 3 (7.5) | 28 (29.2) | 58 (22.1) | 43 (17.6) | 23 (11.4) | 14 (4.8) | 7 (1.6) | 6 (0.7) | 2 (0.2) | 4 (0.7) | 476 (10.3) |
| Skin ulcer | 153 (4.2) | 1 (3.3) | 0 (0.0) | 0 (0.0) | 0 (0.0) | 2 (1.5) | 9 (3.7) | 13 (3.3) | 43 (5.3) | 51 (4.1) | 34 (6.2) | 982 (21.2) |
| Seizure | 152 (3.8) | 5 (11.6) | 3 (6.2) | 3 (2.7) | 6 (3.9) | 11 (6.7) | 16 (6.1) | 22 (5.1) | 34 (3.9) | 39 (3.0) | 13 (2.3) | 659 (14.3) |
| Altered sense of taste | 101 (3.3) | 1 (4.5) | 10 (12.0) | 27 (13.0) | 15 (8.1) | 10 (6.8) | 3 (1.4) | 7 (2.2) | 11 (2.0) | 13 (1.5) | 4 (1.0) | 1602 (34.7) |
| Rash | 78 (1.8) | 5 (11.6) | 3 (3.0) | 2 (0.8) | 1 (0.4) | 4 (2.0) | 5 (1.7) | 17 (3.8) | 18 (2.2) | 15 (1.2) | 8 (1.5) | 384 (8.3) |
| Wheeze | 53 (1.2) | 0 (0.0) | 0 (0.0) | 1 (0.4) | 1 (0.4) | 0 (0.0) | 3 (1.0) | 6 (1.3) | 8 (0.9) | 22 (1.7) | 12 (2.1) | 296 (6.4) |

\* Data are number (percent of patients with non-missing data within columns). Symptoms experienced by <25 individuals are omitted.

In the logistic regression models (Figure 4), similar associations between age and symptoms were seen after adjustment for sex and relationship of onset to admission date as in the unadjusted bar charts. Confusion increased with age. Nausea and vomiting, headache, abdominal pain, and sore throat were each more frequent in younger age groups, decreasing with age. The 95% CI for sex excluded one for most symptoms, but for most the point estimate was close to one. Male patients had greater odds of fever, cough and shortness of breath, and lower odds of gastrointestinal symptoms of nausea and vomiting, diarrhoea, abdominal pain, chest pain, headache and sore throat. The median odds ratio for heterogeneity between countries was greater than the relationship with sex in all symptoms. It was of similar magnitude to the relationship with age for most symptoms. For each symptom, heterogeneity between countries was of a similar magnitude to the effect of age, and a greater magnitude than sex.

**Figure 4.**
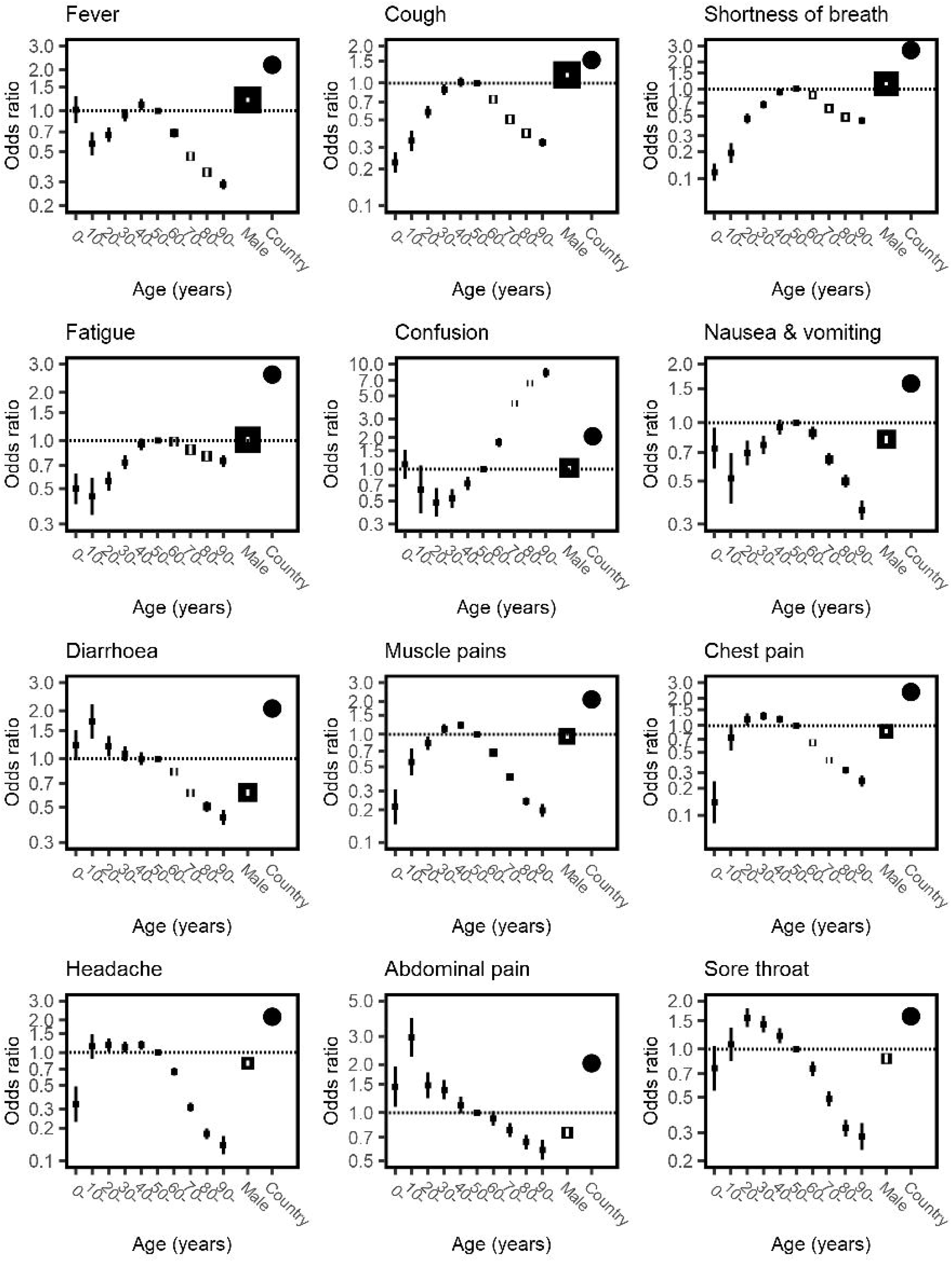
Odds of symptoms among patients admitted to hospital with COVID-19, stratified by age and sex. Each plot is the result of a logistic regression with a symptom as an outcome. Fixed effects of age in ten-year bands (baseline group 50–60 years) and sex are shown in black boxes with 95% confidence intervals. The size of each square is inversely proportional to the variance of the log odds ratio, so larger boxes indicate greater certainty. Clustering by country is included as a random intercept and heterogeneity is depicted by circles showing the median odds ratio.

**Figure 5.**
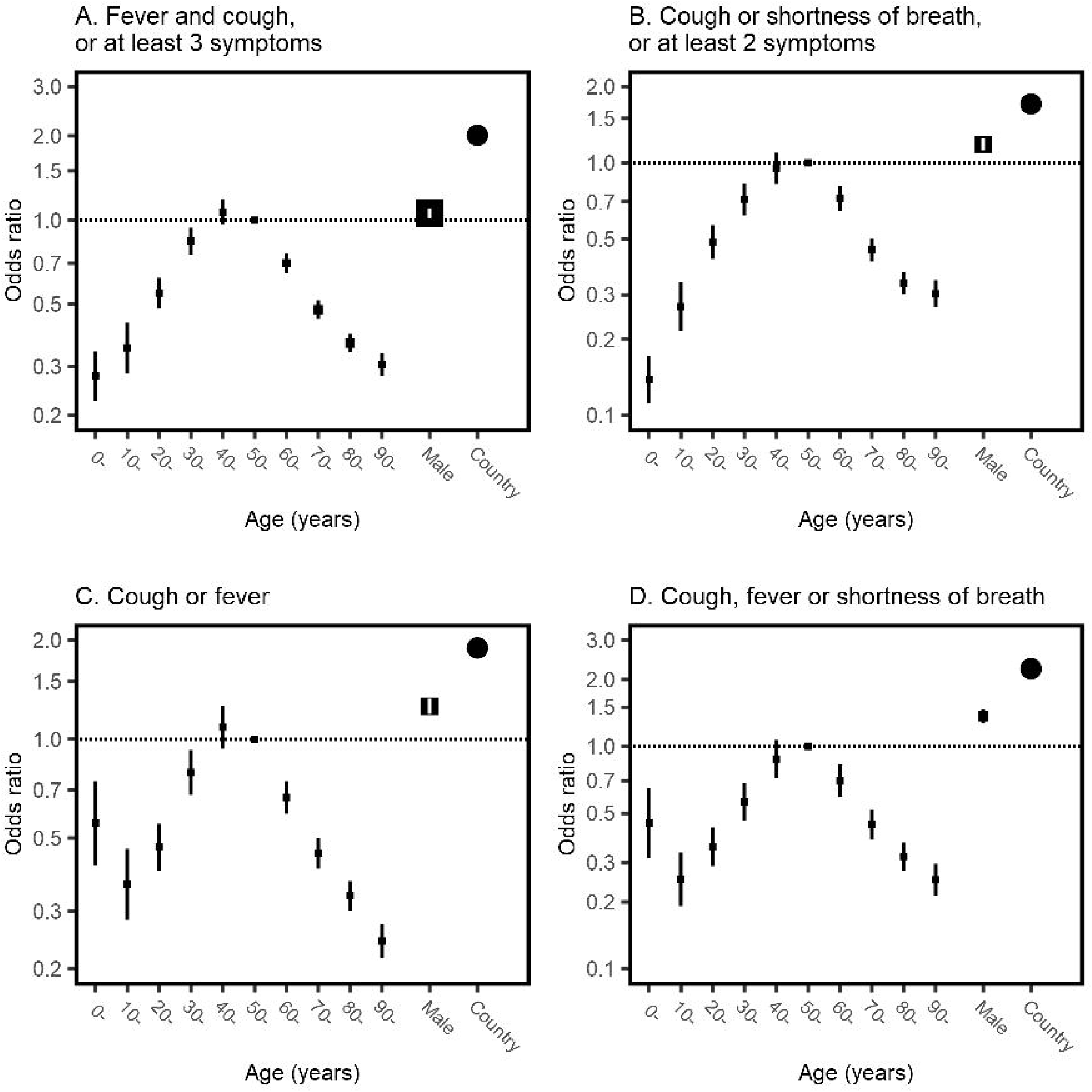
Age- and sex- specific odds of meeting clinical definitions among patients admitted to hospital with COVID-19, stratified by age and sex. Each plot is the result of a logistic regression with a composite group of symptoms as an outcome. Fixed effects of age in ten-year bands (baseline group 50–60 years) and sex are shown in black boxes with 95% confidence intervals. The size of each square is inversely proportional to the variance of the log odds ratio, so larger boxes indicate greater certainty. Clustering by country is included as a random intercept and heterogeneity is depicted by circles showing the median odds ratio. In panel A, the three symptoms are from the list of fever, cough, fatigue, headache, myalgia, sore throat, rhinorrhoea, shortness of breath, nausea and vomiting, diarrhoea, and confusion; in panel B the two symptoms are from the list fever, myalgia, headache, and sore throat. Patients with missing data for cough, fever or shortness of breath are excluded from all four plots.

47⍰280 (79%) patients were included from the United Kingdom. Excluding these patients, the patterns of symptoms were similar to the main analysis (Supplementary Figure 2). The peak prevalence of fever, cough and shortness of breath was in 70-to 80-year-olds, and fatigue increased with age. Below the age of 50 years, the clinical case definitions tended to be less sensitive in the analysis excluding the United Kingdom than in the analysis including it; above the age of 70 years each tended to be more sensitive (grey lines in Figure 3). Within countries, the baseline prevalence of each symptom varied but patterns within countries were broadly similar to the overall results (Supplementary figures 3–14).

## Discussion

The ISARIC prospective multinational cohort study is the largest cohort of patients admitted to hospital with COVID-19. In this report, we confirmed a relationship between patients’ symptoms and their age and sex. The ‘typical’ COVID-19 symptoms occur most frequently in adults aged 30 to 60 years. Commonly used case definitions in use can miss up to half of children and a third of adults over 70 years who are admitted to hospital with COVID-19.

Our results support the findings of smaller cohort studies that atypical symptoms are more common in older adults [21], and correlate with similar findings of atypical presentations for pneumonia, bacteraemia and coronary artery disease [22, 23]. A lower prevalence of symptoms in children and young people has previously been suggested [24, 25], but this is the first large international cohort to collect data prospectively from both adults and children.

Separate analyses of the ISARIC-4C data have identified fever, cough and shortness of breath as frequently co-occurring clusters of symptoms [26, 27]. In children, the next most frequent cluster consisted of systemic, enteric and mucocutaneous symptoms [26]. For adults, other clusters consisted of non-specific viral symptoms, gastrointestinal symptoms, upper respiratory symptoms, neurological symptoms, and symptoms of bronchospasm [27]. Those data were included in this global dataset so the results of these analyses are not independent of our results.

We found that differences in symptoms by sex were statistically significant but generally of smaller magnitude. Typical symptoms of fever, cough and shortness of breath were more common in men than in women; all other symptoms were equal or more common in female patients. A cohort of non-hospitalized patients with COVID-19 in Poland found greater differences in symptoms of lack of appetite (55% of women, 36% of men) and taste disorder (53% women, 40% men) [28]. We are unable to determine from our data whether these differences reflect differences in health-seeking behaviour between men and women, or a biological difference in their response to the infection. Elaboration of this difference should be a goal of future research.

Our results suggest considerable heterogeneity among countries. We have not attempted to elicit reasons for heterogeneity. Potential reasons include cultural idiosyncrasies in reporting symptoms, and hospitals’ criteria for admission and testing. It might also reflect differences in local patient recruitment. Researchers in some countries may be unwilling to recruit confused patients due to requirements for consent, whereas in others the requirement for consent has been waived or could be obtained from a proxy.

We explored the effect of using the country of recruitment as a random effect in regression models and by repeating the analysis excluding the largest country. Each analysis suggested an underlying pattern of lower frequencies of typical symptoms in children and older adults. Therefore, although the prevalence of each symptom reported in this study may not apply to all settings, we have evidence to support the possibility of age-dependent differences internationally.

The size of this cohort is a strength. To our knowledge, it is the largest cohort of hospitalized COVID-19 patients in the world. However, the study has several limitations. Firstly, almost 80% of patients were recruited in a single country. Moreover, less than 1% of patients were recruited from low- or lower-middle-income countries. Secondly, the cohort overwhelmingly includes older adults, with only 1.8% of the cohort aged 18 years or younger. Thirdly, our analysis includes only patients who were hospitalized with COVID-19 and who had a laboratory-confirmed diagnosis. This patient population is more likely to be severely unwell and more likely to exhibit symptoms typically associated with COVID-19 than people who were managed in the community or whose disease has not been recognized. Accordingly, the reporting of ‘typical’ COVID-19 symptoms in this cohort is likely to be an overestimate of the population prevalence. Symptoms are subjective and cannot be externally verified. Some differences for children may reflect that symptoms could only be recorded if a caregiver recognised the symptom or the child had the appropriate vocabulary to describe it. Similarly, some symptoms may be under-reported in elderly patients if there are difficulties in communication, for example due to delirium. As such, the generalizability of estimates of our symptom prevalence is limited. Similarly, there is a shortage of studies conducted outside of high-income countries: a recent scoping review of clinical characteristics of COVID-19 identified no large cohorts in non-high-income countries except China [8].

The absence of a control group of patients without COVID-19 in this dataset prevented estimation of specificity or positive and negative predictive values. We are therefore prevented from advocating changes to clinical case definitions, as such decisions inevitably require a balance of false-positive and false-negative rates. However, given the prevalence of atypical symptoms in our cohort, we can confidently suggest that reliance on clinical case definitions may result in missing cases of COVID-19, especially among children and older adults. Non-healthcare professionals making decisions regarding isolation may be especially vulnerable to missing cases of COVID-19 by adhering to a clinical case definition too strictly.

The reported prevalence of COVID-19 may also rely on a strict interpretation of case definitions. In settings where comprehensive contact tracing is planned, or there is easy access to microbiological testing, a highly sensitive case definition is desirable. However, where decisions are based on clinical diagnoses, it is important to recognize other pathogens that can cause similar constellations of symptoms. The addition of symptoms such as confusion or gastrointestinal symptoms to the COVID-19 case definition could increase sensitivity, but at the cost of reduced specificity. Changes in the senses of taste and smell have recently been added to case definitions. Our data suggest that these criteria would detect only a small proportion of patients admitted to hospital with COVID-19 who were omitted by other definitions.

These results highlight the need to consider COVID-19 even if individuals do not display typical symptoms of the disease. This is especially the case in children and older adults. Given that our results are likely to overestimate the sensitivity of the clinical criteria currently used to identify patients for testing, our results suggest a lower limit to the proportion of people in the community with COVID-19 who would not be identified. Addition of confusion as a symptom would increase the sensitivity of case definitions for older adults; and inclusion of nausea and vomiting or abdominal pain would increase sensitivity for children and young adults. The high proportion of asymptomatic patients identified in patients aged 10 to 20 years suggests that universal screening in these ages could be beneficial when there is widespread community circulation of COVID-19. Ongoing data collection outside high-income countries is needed to establish whether alternative case definitions are needed in different settings. Work is also ongoing to determine whether some constellations of symptoms are associated with better or poorer outcomes than others.

## Data Availability

We welcome applications for UK data through the ISARIC 4C Independent Data and Material Access Committee (https://isaric4c.net). Requests for access to non-UK data can be sent to.

https://www.iddo.org/research-themes/covid-19

## Declarations

### Ethics approval

This observational study required no change to clinical management and permitted enrolment in other research. The study was approved by the World Health Organization Ethics Review Committee (RPC571 and RPC572). Local ethics approval was obtained for each participating country and site according to local requirements.

### Consent to participate

Informed consent was sought where required by local ethics committees. In many jurisdictions, ethics committees and public health authorities approved a waiver of consent to collect anonymised clinical data in the context of this public health emergency.

### Authors’ contributions

Author contributions are listed in the supplementary material.

## Acknowledgements

We are extremely grateful to the frontline clinical and research staff and volunteers, who collected this data in challenging circumstances, and the generosity of the patients and their families for their individual contributions in these difficult times. This work uses data provided by patients and collected by the health institutions and authorities in each country. In the UK, this study involves the National Health Service as part of their care and support #DataSavesLives. We also acknowledge the support of Jeremy J Farrar, Nahoko Shindo, Devika Dixit, Nipunie Rajapakse, Andrew Davison, David Barr, Lyndsey Castle, Martha Buckley, Debbie Malden, Katherine Newell, Kwame O’Neill, Emmanuelle Denis, Claire Petersen, Scott Mullaney, Sue MacFarlane, Nicole Maziere, Julien Martinez, Oslem Dincarslan, Annette Lake and the Irish Critical Care Trials Group. We appreciate the strong collaboration of the WHO Clinical Data Platform Team, including Silvia Bertagnolio, Soe Soe Thwin, and Janet Diaz.

## ISARIC Clinical Characterisation Group

Sheryl Ann Abdukahil, Ryuzo Abe, Laurent Abel, Lara Absil, Andrew Acker, Shingo Adachi, Elisabeth Adam, Diana Adrião, Kate Ainscough, Ali Ait Hssain, Younes Ait Tamlihat, Takako Akimoto, Tala Al-Dabbous, Abdulrahman Al-Fares, Eman Al Qasim, Razi Alalqam, Beatrice Alex, Kévin Alexandre, Huda Alfoudri, Kazali Enagnon Alidjnou, Jeffrey Aliudin, Clotilde Allavena, Nathalie Allou, João Alves, Rita Alves, Maria Amaral, Heidi Ammerlaan, Phoebe Ampaw, Roberto Andini, Claire Andrejak, Andrea Angheben, François Angoulvant, Séverine Ansart, Massimo Antonelli, Carlos Alexandre Antunes De Brito, Yaseen Arabi, Irene Aragao, Antonio Arcadipane, Lukas Arenz, Jean-Benoît Arlet, Christel Arnold-Day, Lovkesh Arora, Elise Artaud-Macari, Angel Asensio, Jean Baptiste Assie, Anika Atique, Johann Auchabie, Hugues Aumaitre, Laurène Azemar, Cécile Azoulay, Benjamin Bach, Delphine Bachelet, J. Kenneth Baillie, Erica Bak, Agamemnon Bakakos, Firouzé Banisadr, Renata Barbalho, Wendy S. Barclay, Michaela Barnikel, Audrey Barrelet, Cleide Barrigoto, Romain Basmaci, Diego Fernando Bautista Rincon, Alexandra Bedossa, Sylvie Behilill, Aleksandr Beljantsev, David Bellemare, Anna Beltrame, Marine Beluze, Nicolas Benech, Dehbia Benkerrou, Suzanne Bennett, LuÍs Bento, Jan-Erik Berdal, Delphine Bergeaud, Lorenzo Bertolino, Simon Bessis, Sybille Bevilcaqua, Krishna Bhavsar, Felwa Bin Humaid, François Bissuel, Patrick Biston, Laurent Bitker, Pablo Blanco-Schweizer, Mathieu Blot, Filomena Boccia, Debby Bogaert, François Bompart, Gareth Booth, Diogo Borges, Raphaël Borie, Jeannet Bos, Hans Martin Bosse, Elisabeth Botelho-Nevers, Lila Bouadma, Olivier Bouchaud, Sabelline Bouchez, Dounia Bouhmani, Damien Bouhour, Kévin Bouiller, Laurence Bouillet, Camille Bouisse, Anne-Sophie Boureau, Maude Bouscambert, Jason Bouziotis, Bianca Boxma, Marielle Boyer-Besseyre, Maria Boylan, Cynthia Braga, Timo Brandenburger, Luca Brazzi, Dorothy Breen, Patrick Breen, Kathy Brickell, Nicolas Brozzi, Nina Buchtele, Christian Buesaquillo, Polina Bugaeva, Marielle Buisson, Erlina Burhan, Ingrid G. Bustos, Denis Butnaru, Sheila Cárcel, André Cabie, Susana Cabral, Eder Caceres, Mia Callahan, Kate Calligy, Jose Andres Calvache, João Camões, Valentine Campana, Paul Campbell, Cecilia Canepa, Mireia Cantero, Pauline Caraux-Paz, Filipa Cardoso, Filipe Cardoso, Sofia Cardoso, Simone Carelli, Nicolas Carlier, Gayle Carney, Chloe Carpenter, Marie-Christine Carret, François Martin Carrier, Gail Carson, Maire-Laure Casanova, Mariana Cascão, José Casimiro, Bailey Cassandra, Silvia Castañeda, Nidyanara Castanheira, Guylaine Castor-Alexandre, Henry Castrillón, Ivo Castro, Ana Catarino, François-Xavier Catherine, Roberta Cavalin, Giulio Giovanni Cavalli, Alexandros Cavayas, Adrian Ceccato, Minerva Cervantes-Gonzalez, Anissa Chair, Catherine Chakveatze, Adrienne Chan, Meera Chand, Julie Chas, Camille Chassin, Anjellica Chen, Yih-Sharng Chen, Matthew Pellan Cheng, Antoine Cheret, Thibault Chiarabini, Julian Chica, Catherine Chirouze, Davide Chiumello, Hwa Jin Cho, Sung Min Cho, Bernard Cholley, Jose Pedro Cidade, Jose Miguel Cisneros Herreros, Barbara Wanjiru Citarella, Anna Ciullo, Jennifer Clarke, Sara Clohisey, Cassidy Codan, Caitriona Cody, Alexandra Coelho, Gwenhaël Colin, Michael Collins, Sebastiano Maria Colombo, Pamela Combs, JP Connelly, Marie Connor, Anne Conrad, Sofía Contreras, Graham S. Cooke, Mary Copland, Hugues Cordel, Amanda Corley, Sarah Cormican, Sabine Cornelis, Arianne Joy Corpuz, Grégory Corvaisier, Camille Couffignal, Sandrine Couffin-Cadiergues, Roxane Courtois, Charles Crepy D’Orleans, Sabine Croonen, Gloria Crowl, Jonathan Crump, Claudina Cruz, Marc Csete, Alberto Cucino, Caroline Cullen, Matthew Cummings, Gerard Curley, Elodie Curlier, Paula Custodio, Frédérick D’Aragon, Ana Da Silva Filipe, Charlene Da Silveira, Eric D’Ortenzio, Al-Awwab Dabaliz, Andrew B. Dagens, Heidi Dalton, Jo Dalton, Nick Daneman, Emmanuelle A. Dankwa, Jorge Dantas, Nathalie De Castro, Diego De Mendoza, Rafael Freitas De Oliveira França, Rosanna De Rosa, Thushan De Silva, Peter De Vries, David Dean, Marie-Pierre Debray, William Dechert, Lauren Deconninck, Romain Decours, Isabelle Delacroix, Karen Delavigne, Ionna Deligiannis, Andrea Dell’amore, Pierre Delobel, Elisa Demonchy, Emmanuelle Denis, Dominique Deplanque, Dominique Deplanque, Pieter Depuydt, Mehul Desai, Diane Descamps, Mathilde Desvallée, Santi Rahayu Dewayanti, Alpha Diallo, Sylvain Diamantis, André Dias, Juan Jose Diaz Diaz, Rodrigo Diaz, Kévin Didier, Jean-Luc Diehl, Wim Dieperink, Jérôme Dimet, Vincent Dinot, Alphonsine Diouf, Yael Dishon, Félix Djossou, Annemarie B. Docherty, Andy Dong, Christl A. Donnelly, Maria Donnelly, Chloe Donohue, Céline Dorival, James Joshua Douglas, Renee Douma, Nathalie Dournon, Triona Downer, Mark Downing, Tom Drake, Vincent Dubee, François Dubos, Alexandra Ducancelle, Susanne Dudman, Jake Dunning, Emanuele Durante Mangoni, Silvia Duranti, Lucian Durham III, Bertrand Dussol, Xavier Duval, Anne Margarita Dyrhol-Riise, Carla Eira, José Ernesto Vidal, Mohammed El Sanharawi, Subbarao Elapavaluru, Brigitte Elharrar, Natalie Elkheir, Jacobien Ellerbroek, Rachael Ellis, Philippine Eloy, Tarek Elshazly, Isabelle Enderle, Ilka Engelmann, Vincent Enouf, Olivier Epaulard, Mariano Esperatti, Hélène Esperou, Marina Esposito-Farese, João Estevão, Manuel Etienne, Manuel Etienne, Nadia Ettalhaoui, Anna Greti Everding, Mirjam Evers, Isabelle Fabre, Amna Faheem, Arabella Fahy, Cameron J. Fairfield, Pedro Faria, Nataly Farshait, Arie Zainul Fatoni, Karine Faure, Mohamed Fayed, Niamh Feely, Jorge Fernandes, Marília Fernandes, Susana Fernandes, Joana Ferrão, Eglantine Ferrand Devouge, Mário Ferraz, Benigno Ferreira, Ricard Ferrer-Roca, Claudia Figueiredo-Mello, Clara Flateau, Tom Fletcher, Letizia Lucia Florio, Claire Foley, Victor Fomin, Claudio Duarte Fonseca, Tatiana Fonseca, Patricia Fontela, Simon Forsyth, Giuseppe Foti, Erwan Fourn, Rob Fowler, Diego Franch-Llasat, Christophe Fraser, John Fraser, Marcela Vieira Freire, Ana Freitas Ribeiro, Caren Friedrich, Stéphane Fry, Nora Fuentes, Masahiro Fukuda, Joan Gómez-Junyent, Valérie Gaborieau, Benoît Gachet, Rostane Gaci, Massimo Gagliardi, Jean-Charles Gagnard, Amandine Gagneux-Brunon, Sérgio Gaião, Phil Gallagher, Elena Gallego Curto, Carrol Gamble, Arthur Garan, Esteban Garcia-Gallo, Rebekha Garcia, Denis Garot, Valérie Garrait, Nathalie Gault, Aisling Gavin, Alexandre Gaymard, Johannes Gebauer, Louis Gerbaud Morlaes, Nuno Germano, Jade Ghosn, Marco Giani, Carlo Giaquinto, Jess Gibson, Tristan Gigante, Morgane Gilg, Guillermo Giordano, Michelle Girvan, Valérie Gissot, Gezy Giwangkancana, Daniel Glikman, Petr Glybochko, Eric Gnall, Geraldine Goco, François Goehringer, Siri Goepel, Jean-Christophe Goffard, Jonathan Golob, Isabelle Gorenne, Cécile Goujard, Tiphaine Goulenok, Margarite Grable, Edward Wilson Grandin, Pascal Granier, Giacomo Grasselli, Christopher A. Green, William Greenhalf, Segolène Greffe, Domenico Luca Grieco, Matthew Griffee, Fiona Griffiths, Ioana Grigoras, Albert Groenendijk, Anja Grosse Lordemann, Heidi Gruner, Yusing Gu, Jérémie Guedj, Dewi Guellec, Anne-Marie Guerguerian, Daniela Guerreiro, Romain Guery, Anne Guillaumot, Laurent Guilleminault, Thomas Guimard, Daniel Haber, Sheeba Hakak, Matthew Hall, Sophie Halpin, Ansley Hamer, Rebecca Hamidfar, Terese Hammond, Hayley Hardwick, Kristen Harley, Ewen M. Harrison, Janet Harrison, Leanne Hays, Jan Heerman, Lars Heggelund, Ross Hendry, Martina Hennessy, Aquiles Henriquez-Trujillo, Maxime Hentzien, Jaime Hernandez-Montfort, Astarini Hidayah, Dawn Higgins, Eibhilin Higgins, Samuel Hinton, Ana Hipólito -Reis, Hiroaki Hiraiwa, Julian A. Hiscox, Antonia Ying Wai Ho, Alexandre Hoctin, Isabelle Hoffmann, Oscar Hoiting, Rebecca Holt, Jan Cato Holter, Peter Horby, Juan Pablo Horcajada, Koji Hoshino, Kota Hoshino, Catherine L. Hough, Jimmy Ming-Yang Hsu, Jean-Sébastien Hulot, Samreen Ijaz, Hajnal-Gabriela Illes, Hugo Inácio, Carmen Infante Dominguez, Elias Iosifidis, Lacey Irvine, Sarah Isgett, Tiago Isidoro, Margaux Isnard, Junji Itai, Daniel Ivulich, Salma Jaafoura, Julien Jabot, Clare Jackson, Nina Jamieson, Stéphane Jaureguiberry, Jeffrey Javidfar, Zabbe Jean-Benoît, Florence Jego, Synne Jenum, Ruth Jimbo Sotomayor, Ruth Noemí Jorge GarcÍa, Cédric Joseph, Mark Joseph, Philippe Jouvet, Hanna Jung, Ouifiya Kafif, Florentia Kaguelidou, Sabina Kali, Smaragdi Kalomoiri, Darshana Hewa Kandamby, Chris Kandel, Ravi Kant, Christiana Kartsonaki, Daisuke Kasugai, Kevin Katz, Simreen Kaur Johal, Sean Keating, Andrea Kelly, Sadie Kelly, Lisa Kennedy, Kalynn Kennon, Younes Kerroumi, Evelyne Kestelyn, Imrana Khalid, Antoine Khalil, Coralie Khan, Irfan Khan, Michelle E Kho, Saye Khoo, Yuri Kida, Peter Kiiza, Anders Benjamin Kildal, Antoine Kimmoun, Detlef Kindgen-Milles, Nobuya Kitamura, Paul Klenerman, Gry Kloumann Bekken, Stephen Knight, Robin Kobbe, Malte Kohns Vasconcelos, Volkan Korten, Caroline Kosgei, Karolina Krawczyk, Pavan Kumar Vecham, Deepali Kumar, Ethan Kurtzman, Demetrios Kutsogiannis, Konstantinos Kyriakoulis, Erwan L’her, Marie Lachatre, Marie Lacoste, John G Laffey, Marie Lagrange, Fabrice Laine, Marc Lambert, François Lamontagne, Marie Langelot-Richard, Eka Yudha Lantang, Marina Lanza, Cédric Laouénan, Samira Laribi, Delphine Lariviere, Odile Launay, Yoan Lavie-Badie, Andrew Law, Clément Le Bihan, Cyril Le Bris, Eve Le Coustumier, Georges Le Falher, Sylvie Le Gac, Quentin Le Hingrat, Marion Le Maréchal, Soizic Le Mestre, Vincent Le Moing, Hervé Le Nagard, Paul Le Turnier, Rafael León, Minh Le, Marta Leal Santos, Ema Leal, James Lee, Su Hwan Lee, Todd Lee, Gary Leeming, Bénédicte Lefebvre, Laurent Lefebvre, Benjamin Lefevre, François Lellouche, Adrien Lemaignen, Véronique Lemee, Gretchen Lemmink, Michela Leone, Quentin Lepiller, François-Xavier Lescure, Olivier Lesens, Mathieu Lesouhaitier, Claire Levy-Marchal, Bruno Levy, Yves Levy, Gianluigi Li Bassi, Janet Liang, Wei Shen Lim, Bruno Lina, Andreas Lind, Guillaume Lingas, Sylvie Lion-Daolio, Keibun Liu, Antonio Loforte, Navy Lolong, Diogo Lopes, Dalia Lopez-Colon, Paul Loubet, Jean Christophe Lucet, Carlos M. Luna, Olguta Lungu, Liem Luong, Dominique Luton, Ruth Lyons, Fredrik Müller, Karl Erik Müller, Olavi Maasikas, Sarah Macdonald, Moïse Machado, Gabrielle Macheda, Juan Macias Sanchez, Jai Madhok, Rafael Mahieu, Sophie Mahy, Lars Siegfrid Maier, Mylène Maillet, Thomas Maitre, Maximilian Malfertheiner, Nadia Malik, Fernando Maltez, Denis Malvy, Marina Mambert, Victoria Manda, Jose M. Mandei, Edmund Manning, Aldric Manuel, Ceila Maria Sant’Ana Malaque, Flávio Marino, Carolline De Araújo Mariz, Charbel Maroun Eid, Ana Marques, Catherine Marquis, Brian Marsh, Laura Marsh, John Marshall, Celina Turchi Martelli, Guillaume Martin-Blondel, Ignacio Martin-Loeches, Alejandro Martin-Quiros, Dori-Ann Martin, Emily Martin, Martin Martinot, Caroline Martins Rego, Ana Martins, João Martins, Gennaro Martucci, Eva Miranda Marwali, Juan Fernado Masa Jimenez, David Maslove, Sabina Mason, Moshe Matan, Daniel Mathieu, Mathieu Mattei, Romans Matulevics, Laurence Maulin, Natalie Mc Evoy, Aine McCarthy, Anne McCarthy, Colin McCloskey, Rachael McConnochie, Sherry McDermott, Sarah McDonald, Samuel McElwee, Natalie McEvoy, Allison McGeer, Niki McGuinness, Kenneth A. McLean, Bairbre McNicholas, Edel Meaney, Cécile Mear-Passard, Maggie Mechlin, Ferruccio Mele, Kusum Menon, France Mentré, Alexander J. Mentzer, Noémie Mercier, Antoine Merckx, Blake Mergler, Laura Merson, António Mesquita, Agnès Meybeck, Alison M. Meynert, Vanina Meyssonnier, Amina Meziane, Medhi Mezidi, Céline Michelanglei, Vladislav Mihnovitš, Hugo Miranda Maldonado, Mary Mone, Asma Moin, David Molina, Elena Molinos, Agostinho Monteiro, Claudia Montes, Giorgia Montrucchio, Sarah Moore, Shona C. Moore, Lina Morales-Cely, Lucia Moro, Diego Rolando Morocho Tutillo, Ana Motos, Hugo Mouquet, Clara Mouton Perrot, Julien Moyet, Jimmy Mullaert, Daniel Munblit, Derek Murphy, Marlène Murris, Dimitra Melia Myrodia, Yohan N’guyen, Nadège Neant, Holger Neb, Nikita A Nekliudov, Raul Neto, Emily Neumann, Bernardo Neves, Pauline Yeung Ng, Wing Yiu Ng, Orna Ni Choileain, Alistair Nichol, Stephanie Nonas, Marion Noret, Lisa Norman, Alessandra Notari, Mahdad Noursadeghi, Karolina Nowicka, Saad Nseir, Jose I Nunez, Elsa Nyamankolly, Max O’Donnell, Katie O’Hearn, Conar O’Neil, Giovanna Occhipinti, Tawnya Ogston, Takayuki Ogura, Tak-Hyuk Oh, Shinichiro Ohshimo, Budha Charan Singh Oinam, Ana Pinho Oliveira, João Oliveira, Piero Olliaro, David S. Y. Ong, Wilna Oosthuyzen, Peter J.M. Openshaw, Claudia Milena Orozco-Chamorro, Andrés Orquera, Javier Osatnik, Nadia Ouamara, Rachida Ouissa, Clark Owyang, Eric Oziol, Diana Póvoas, Maïder Pagadoy, Justine Pages, Mario Palacios, Massimo Palmarini, Giovanna Panarello, Prasan Kumar Panda, Mauro Panigada, Nathalie Pansu, Aurélie Papadopoulos, Briseida Parra, Jérémie Pasquier, Fabian Patauner, Luís Patrão, Christelle Paul, Mical Paul, Jorge Paulos, William A. Paxton, Jean-François Payen, India Pearse, Giles J Peek, Florent Peelman, Nathan Peiffer-Smadja, Vincent Peigne, Mare Pejkovska, Ithan D. Peltan, Rui Pereira, Daniel Perez, Thomas Perpoint, Antonio Pesenti, Lenka Petroušová, Ventzislava Petrov-Sanchez, Gilles Peytavin, Scott Pharand, Michael Piagnerelli, Walter Picard, Olivier Picone, Marie Piel-Julian, Carola Pierobon, Carlos Pimentel, Lionel Piroth, Riinu Pius, Simone Piva, Laurent Plantier, Daniel Plotkin, Julien Poissy, Maria Pokorska-Spiewak, Sergio Poli, Georgios Pollakis, Jolanta Popielska, Douwe F. Postma, Pedro Povoa, Jeff Powis, Sofia Prapa, Christian Prebensen, Jean-Charles Preiser, Vincent Prestre, Nicholas Price, Anton Prinssen, Mark G. Pritchard, Lúcia Proença, Oriane Puéchal, Gregory Purcell, Luisa Quesada, Else Quist-Paulsen, Mohammed Quraishi, Indrek Rätsep, Bernhard Rössler, Christian Rabaud, Marie Rafiq, Gabrielle Ragazzo, Fernando Rainieri, Nagarajan Ramakrishnan, Kollengode Ramanathan, Blandine Rammaert, Christophe Rapp, Menaldi Rasmin, Cornelius Rau, Stanislas Rebaudet, Sarah Redl, Brenda Reeve, Liadain Reid, Renato Reis, Jonathan Remppis, Martine Remy, Hanna Renk, Liliana Resende, Anne-Sophie Resseguier, Matthieu Revest, Oleksa Rewa, Luis Felipe Reyes, David Richardson, Denise Richardson, Laurent Richier, Jordi Riera, Ana Lúcia Rios, Asgar Rishu, Patrick Rispal, Karine Risso, Maria Angelica Rivera Nuñez, Nicholas Rizer, André Roberto, Stephanie Roberts, David L. Robertson, Olivier Robineau, Ferran Roche-Campo, Paola Rodari, Simão Rodeia, Julia Rodriguez Abreu, Emmanuel Roilides, Amanda Rojek, Juliette Romaru, Roberto Roncon-Albuquerque Jr, Mélanie Roriz, Manuel Rosa-Calatrava, Michael Rose, Dorothea Rosenberger, Andrea Rossanese, Bénédicte Rossignol, Patrick Rossignol, Carine Roy, Benoît Roze, Clark D. Russell, Steffi Ryckaert, Aleksander Rygh Holten, Xavier Sánchez Choez, Isabela Saba, Musharaf Sadat, Nadia Saidani, Leonardo Salazar, Gabriele Sales, Stéphane Sallaberry, Hélène Salvator, Angel Sanchez-Miralles, Olivier Sanchez, Vanessa Sancho-Shimizu, Gyan Sandhu, Oana Sandulescu, Marlene Santos, Shirley Sarfo-Mensah, Benjamine Sarton, Egle Saviciute, Parthena Savvidou, Joshua Scarsbrook, Tjard Schermer, Arnaud Scherpereel, Marion Schneider, Stephan Schroll, Michael Schwameis, James Scott-Brown, Janet T. Scott, Nicholas Sedillot, Tamara Seitz, Caroline Semaille, Malcolm G. Semple, Eric Senneville, Filipa Sequeira, Tânia Sequeira, Ellen Shadowitz, Mohammad Shamsah, Pratima Sharma, Catherine A. Shaw, Victoria Shaw, Nisreen Shiban, Nobuaki Shime, Hiroaki Shimizu, Keiki Shimizu, Sally Shrapnel, Hoi Ping Shum, Nassima Si Mohammed, Louise Sigfrid, Catarina Silva, Maria Joao Silva, Wai Ching Sin, Vegard Skogen, Sue Smith, Benjamin Smood, Michelle Smyth, Morgane Snacken, Dominic So, Monserrat Solis, Joshua Solomon, Tom Solomon, Emily Somers, Agnès Sommet, Myung Jin Song, Rima Song, Tae Song, Michael Sonntagbauer, Edouard Soum, Maria Sousa Uva, Marta Sousa, Vicente Souza-Dantas, Alexandra Sperry, Shiranee Sriskandan, Thomas Staudinger, Stephanie-Susanne Stecher, Ymkje Stienstra, Birgitte Stiksrud, Adrian Streinu-Cercel, Anca Streinu-Cercel, Samantha Strudwick, Ami Stuart, David Stuart, Asfia Sultana, Charlotte Summers, Magdalena Surovcová Andrey A Svistunov, Konstantinos Syrigos, Jaques Sztajnbok, Konstanty Szuldrzynski, François Téoulé, Shirin Tabrizi, Lysa Tagherset, Ewa Talarek, Sara Taleb, Jelmer Talsma, Le Van Tan, Hiroyuki Tanaka, Taku Tanaka, Hayato Taniguchi, Coralie Tardivon, Pierre Tattevin, M Azhari Taufik, Richard S. Tedder, João Teixeira, Marie-Capucine Tellier, Pleun Terpstra, Olivier Terrier, Nicolas Terzi, Hubert Tessier-Grenier, Vincent Thibault, Simon-Djamel Thiberville, Benoît Thill, A.A. Roger Thompson, Shaun Thompson, David Thomson, Emma C. Thomson, Duong Bich Thuy, Ryan S. Thwaites, Peter S. Timashev, Jean-François Timsit, Bharath Kumar Tirupakuzhi Vijayaraghavan, Maria Toki, Kristian Tonby, Rosario Maria Torres Santos-Olmo, Antoni Torres, Margarida Torres, Théo Trioux, Huynh Trung Trieu, Cécile Tromeur, Ioannis Trontzas, Jonathan Troost, Tiffany Trouillon, Christelle Tual, Sarah Tubiana, Helen Tuite, Lance C.W. Turtle, Pawel Twardowski, Makoto Uchiyama, Roman Ullrich, Alberto Uribe, Asad Usman, Luís Val-Flores, Stijn Van De Velde, Marcel Van Den Berge, Machteld Van Der Feltz, Nicky Van Der Vekens, Peter Van Der Voort, Sylvie Van Der Werf, Marlice Van Dyk, Laura Van Gulik, Jarne Van Hattem, Steven Van Lelyveld, Carolien Van Netten, Noémie Vanel, Henk Vanoverschelde, Charline Vauchy, Aurélie Veislinger, Jorge Velazco, Sara Ventura, Annelies Verbon, César Vieira, Joy Ann Villanueva, Judit Villar, Pierre-Marc Villeneuve, Andrea Villoldo, Nguyen Van Vinh Chau, Benoit Visseaux, Hannah Visser, Aapeli Vuorinen, Fanny Vuotto, Chih-Hsien Wang, Jia Wei, Katharina Weil, Sanne Wesselius, Murray Wham, Bryan Whelan, Nicole White, Aurélie Wiedemann, Keith Wille, Evert-Jan Wils, Ioannis Xynogalas, Jacky Y Suen, Sophie Yacoub, Masaki Yamazaki, Yazdan Yazdanpanah, Cécile Yelnik, Stephanie Yerkovich, Toshiki Yokoyama, Hodane Yonis, Paul Young, Saptadi Yuliarto, Marion Zabbe, Kai Zacharowski, Maram Zahran, Maria Zambon, Alberto Zanella, Konrad Zawadka, Hiba Zayyad, Alexander Zoufaly, David Zucman; The PETAL Network Investigators, The Western Australian Covid-19 Research Response.

