## Supplementary material for "COVID-19 symptoms at hospital admission vary with age and sex: ISARIC multinational study"

ISARIC Covid-19 Collaboration

**Supplementary Table 1. Patients and sites included in this analysis**

| **Country** |  | **Participants*** | **Sites** |
| --- | --- | --- | --- |
| *East Asia & Pacific* | |  |  |
|  | China | 11 (0.0) | 2 |
|  | Hong Kong SAR, China | 18 (0.0) | 2 |
|  | Indonesia | 76 (0.1) | 5 |
|  | Japan | 46 (0.1) | 11 |
|  | Korea South | 4 (0.0) | 4 |
|  | Malaysia | 3087 (5.1) | 1 |
|  | New Zealand | 9 (0.0) | 6 |
|  | Taiwan | 1 (0.0) | 1 |
| *Europe & Central Asia* | |  |  |
|  | Austria | 11 (0.0) | 2 |
|  | Belgium | 277 (0.5) | 3 |
|  | Czech Republic | 2 (0.0) | 1 |
|  | Estonia | 23 (0.0) | 2 |
|  | France | 2979 (5.0) | 2 |
|  | Germany | 82 (0.1) | 7 |
|  | Greece | 32 (0.1) | 1 |
|  | Ireland | 510 (0.8) | 13 |
|  | Italy | 333 (0.6) | 9 |
|  | Netherlands | 859 (1.4) | 13 |
|  | Norway | 191 (0.3) | 4 |
|  | Poland | 13 (0.0) | 2 |
|  | Portugal | 219 (0.4) | 12 |
|  | Romania | 543 (0.9) | 1 |
|  | Russian Federation | 1662 (2.8) | 1 |
|  | Spain | 311 (0.5) | 11 |
|  | United Kingdom | 47332 (78.8) | 209 |
| *Latin America & Caribbean* | |  |  |
|  | Argentina | 4 (0.0) | 2 |
|  | Brazil | 12 (0.0) | 2 |
|  | Chile | 2 (0.0) | 1 |
|  | Colombia | 112 (0.2) | 3 |
|  | Ecuador | 1 (0.0) | 1 |
|  | Mexico | 2 (0.0) | 1 |
|  | Peru | 28 (0.0) | 1 |
|  | St. Vincent and the Grenadines | 1 (0.0) | 1 |
| *Middle East & North Africa* | |  |  |
|  | Israel | 72 (0.1) | 2 |
|  | Kuwait | 18 (0.0) | 2 |
|  | Qatar | 1 (0.0) | 1 |
| *North America* | |  |  |
|  | Canada | 420 (0.7) | 19 |
|  | United States | 500 (0.8) | 26 |
| *South Asia* | |  |  |
|  | India | 267 (0.4) | 3 |
| *Sub-Saharan Africa* | |  |  |
|  | Congo | 1 (0.0) | 1 |
|  | Ghana | 1 (0.0) | 1 |
|  | Sao Tome and Principe | 1 (0.0) | 1 |
|  | South Africa | 7 (0.0) | 1 |
| * Number of patients included in this analysis (percent of all patients in the analysis) | | | |

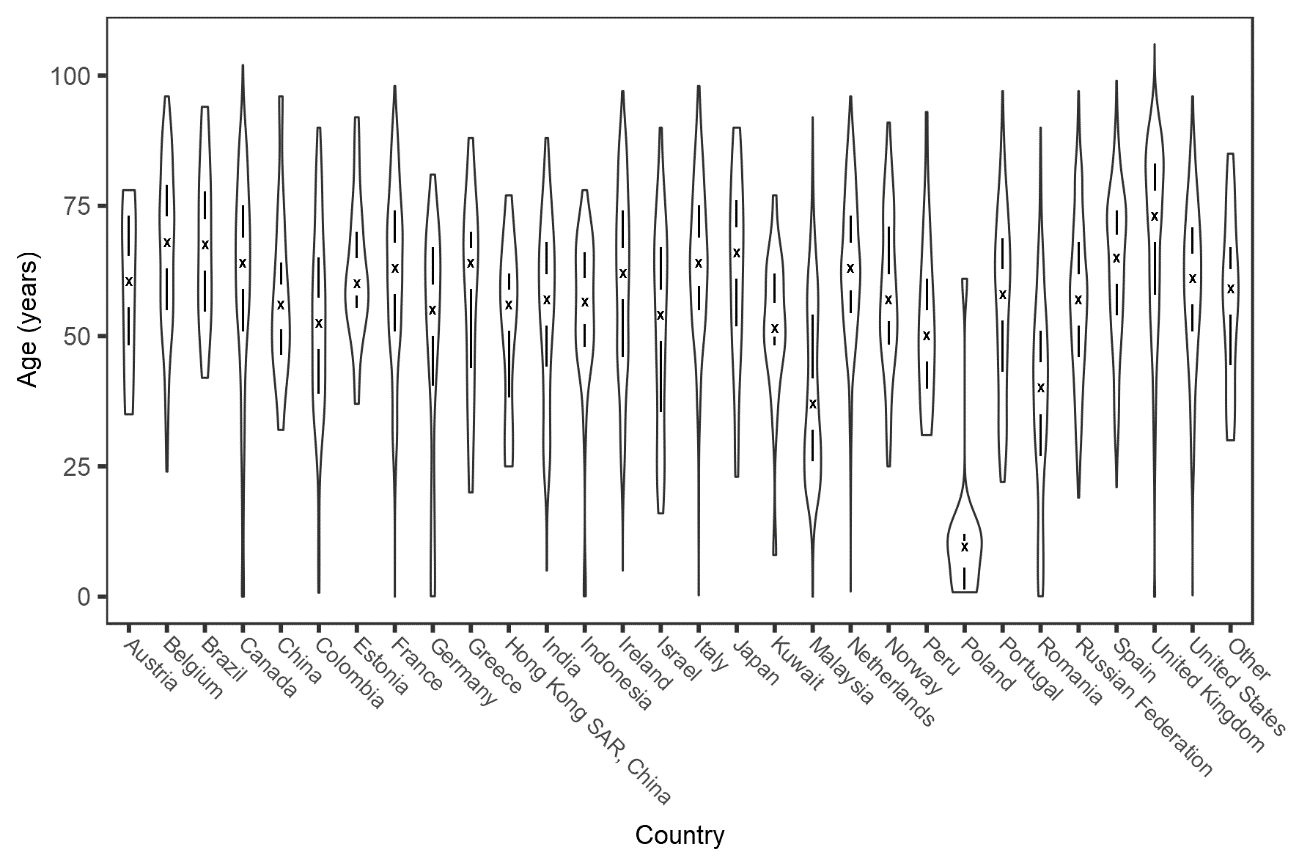

**Supplementary Figure 1. Violin plot of age distribution of patients recruited by country.**

The outline area shows the age distribution within each country. The cross marks the median age and the vertical lines mark the interquartile range. Countries with fewer than ten patients are included as ‘Other’.

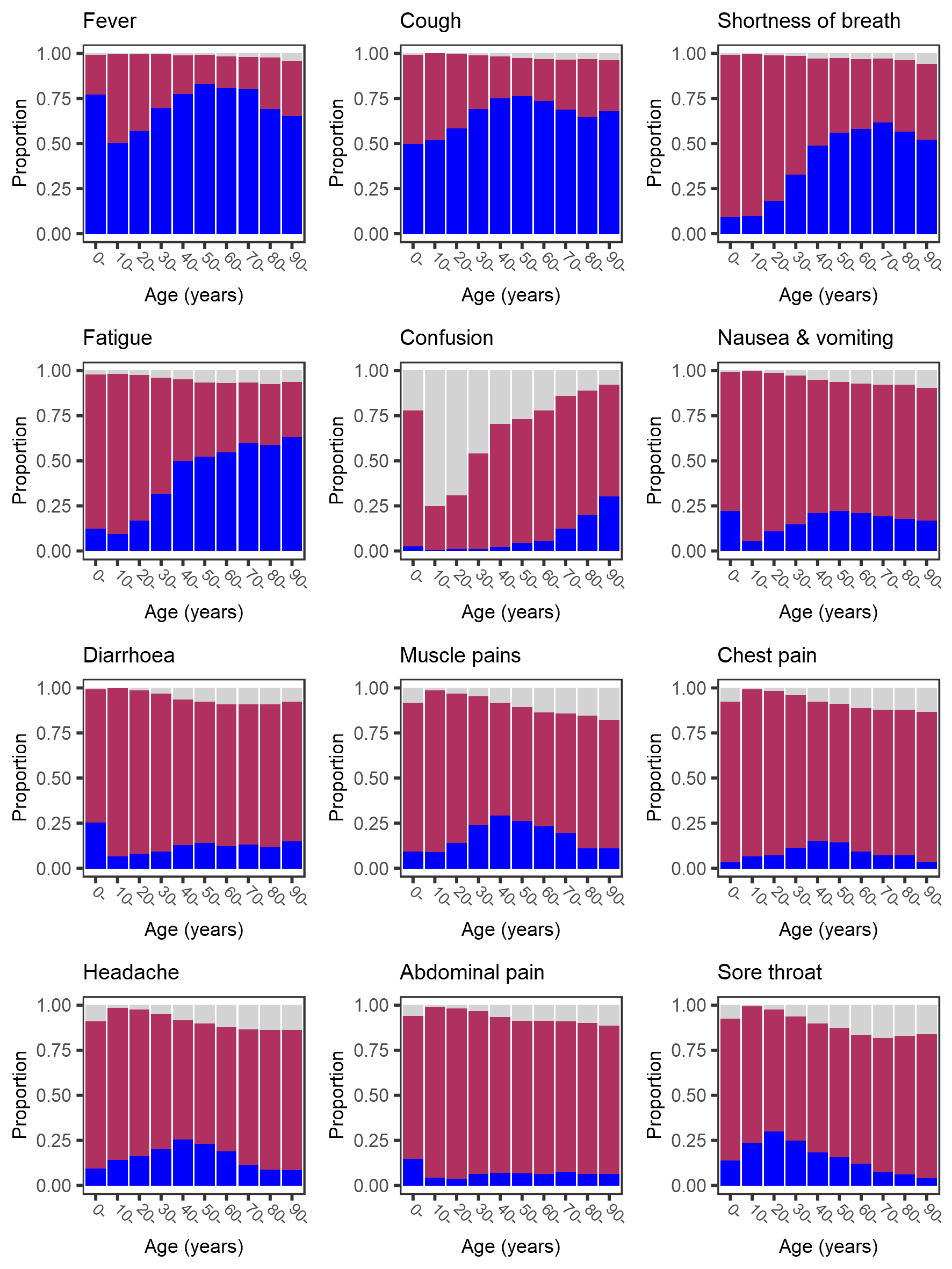

**Supplementary Figure 3. Age-specific prevalence of symptoms at hospital admission with covid‑19, excluding patients recruited in the United Kingdom.**

Dark blue bars show symptom present, maroon bars show symptom absent, pale grey bars show missing data.

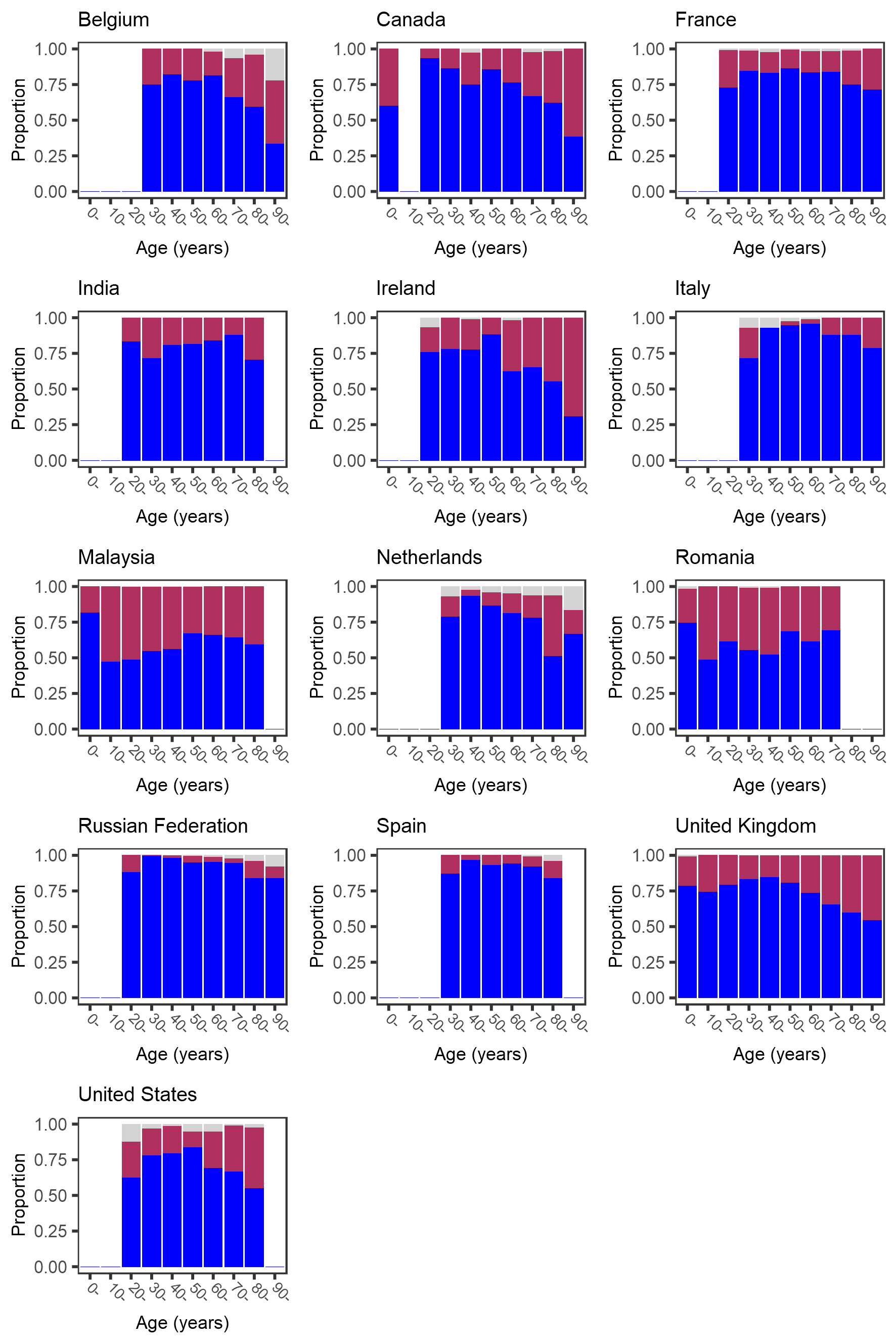

**Supplementary Figure 3. Age-specific prevalence of fever at hospital admission with covid‑19.**

Dark blue bars show symptom present, maroon bars show symptom absent, pale grey bars show missing data. Bars that would represent <10 individuals are omitted.

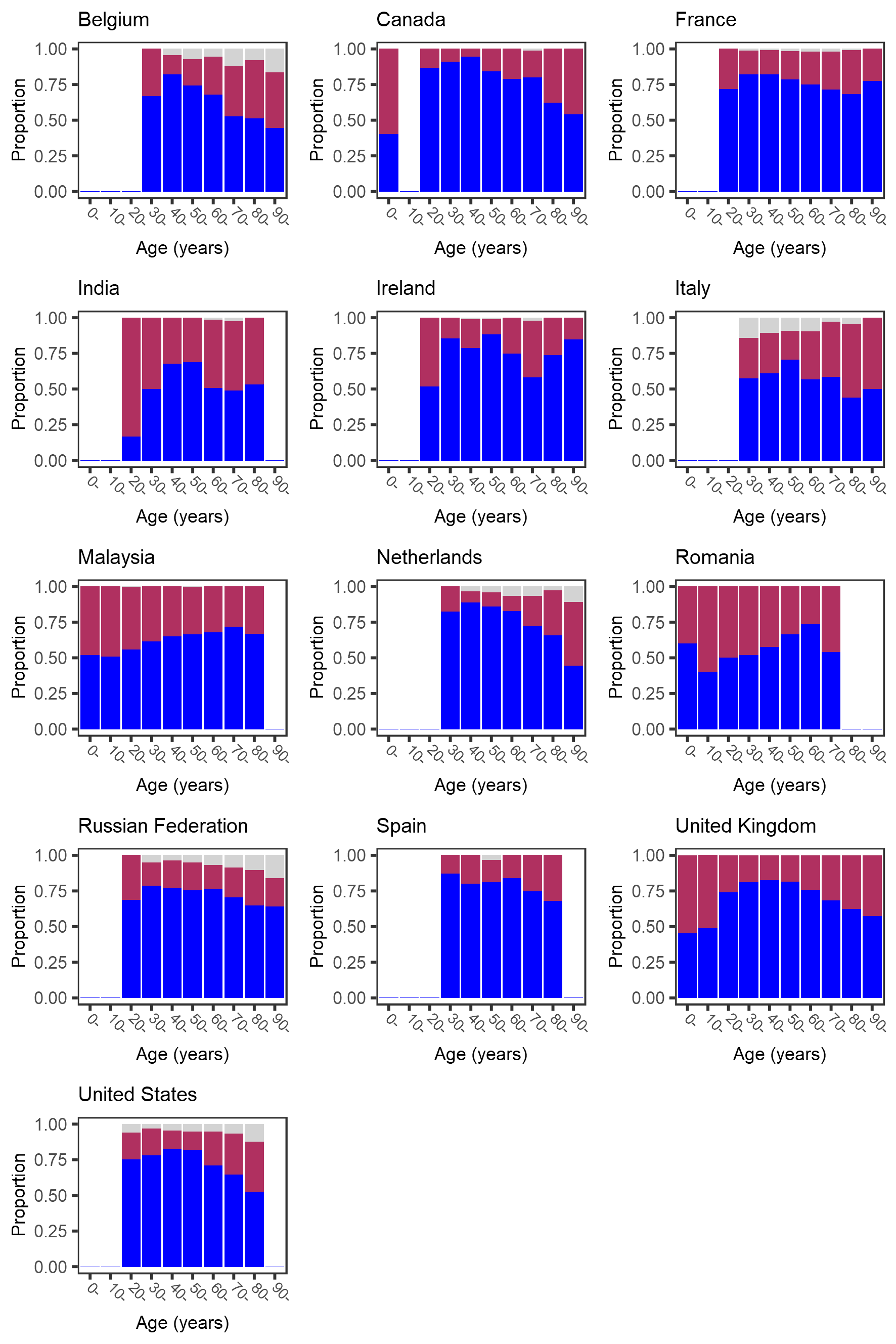

**Supplementary Figure 4. Age-specific prevalence of cough at hospital admission with covid‑19.**

Dark blue bars show symptom present, maroon bars show symptom absent, pale grey bars show missing data. Bars that would represent <10 individuals are omitted.

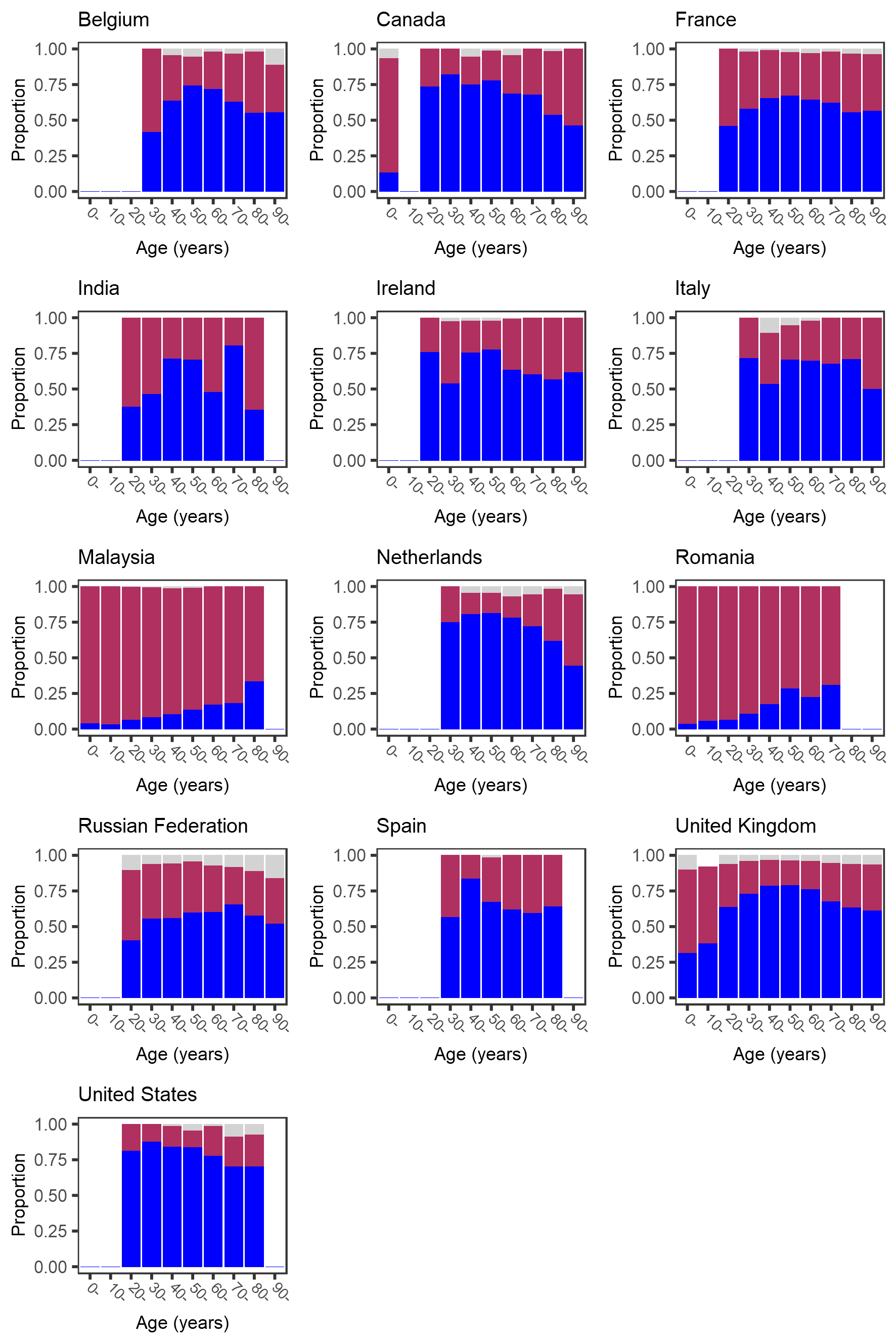

**Supplementary Figure 5. Age-specific prevalence of shortness of breath at hospital admission with covid‑19.** Dark blue bars show symptom present, maroon bars show symptom absent, pale grey bars show missing data. Bars that would represent <10 individuals are omitted.

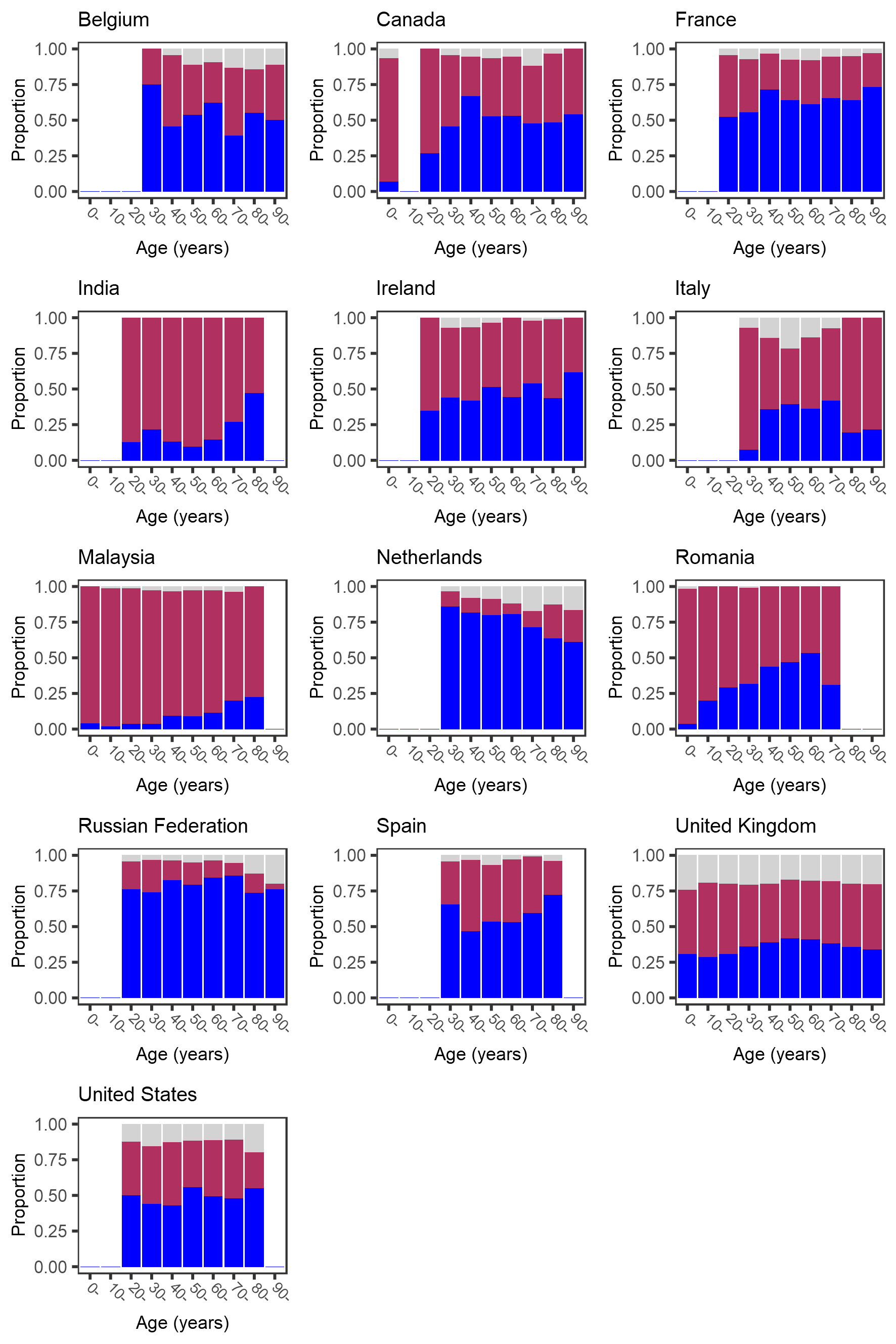

**Supplementary Figure 6. Age-specific prevalence of fatigue at hospital admission with covid‑19.**

Dark blue bars show symptom present, maroon bars show symptom absent, pale grey bars show missing data. Bars that would represent <10 individuals are omitted.

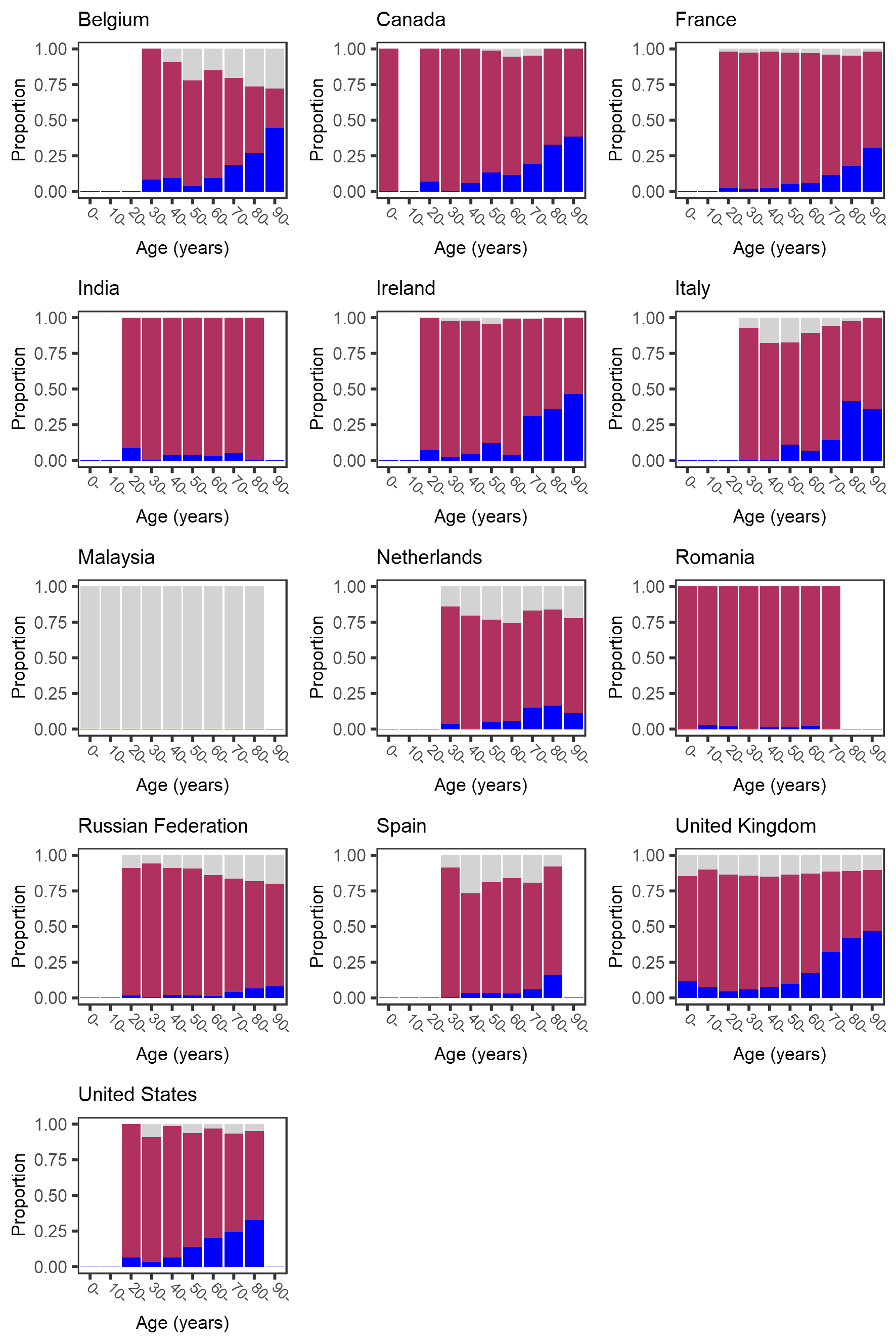

**Supplementary Figure 7. Age-specific prevalence of confusion at hospital admission with covid‑19.**

Dark blue bars show symptom present, maroon bars show symptom absent, pale grey bars show missing data. Bars that would represent <10 individuals are omitted.

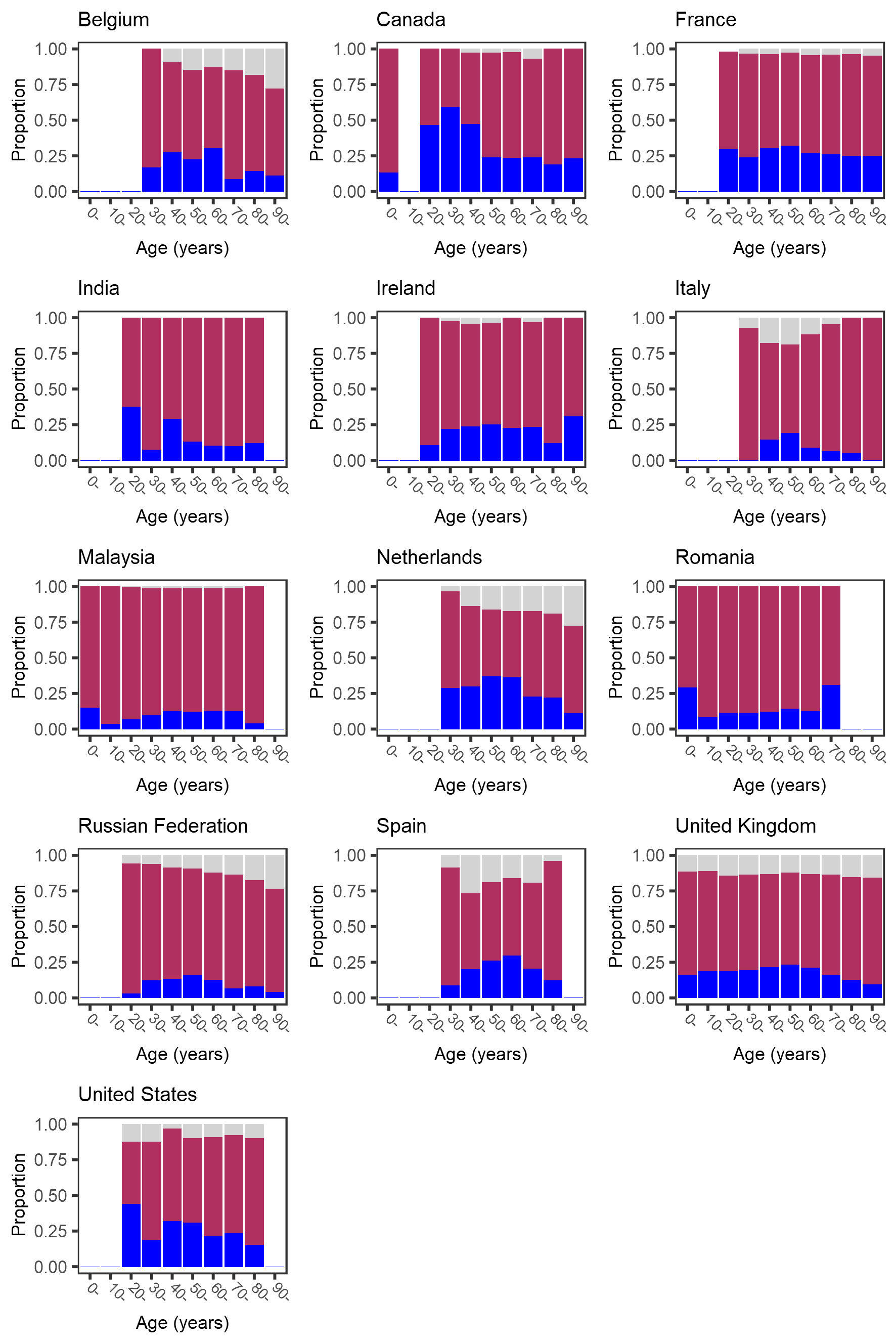

**Supplementary Figure 8. Age-specific prevalence of nausea and vomiting at hospital admission with covid‑19.** Dark blue bars show symptom present, maroon bars show symptom absent, pale grey bars show missing data. Bars that would represent <10 individuals are omitted.

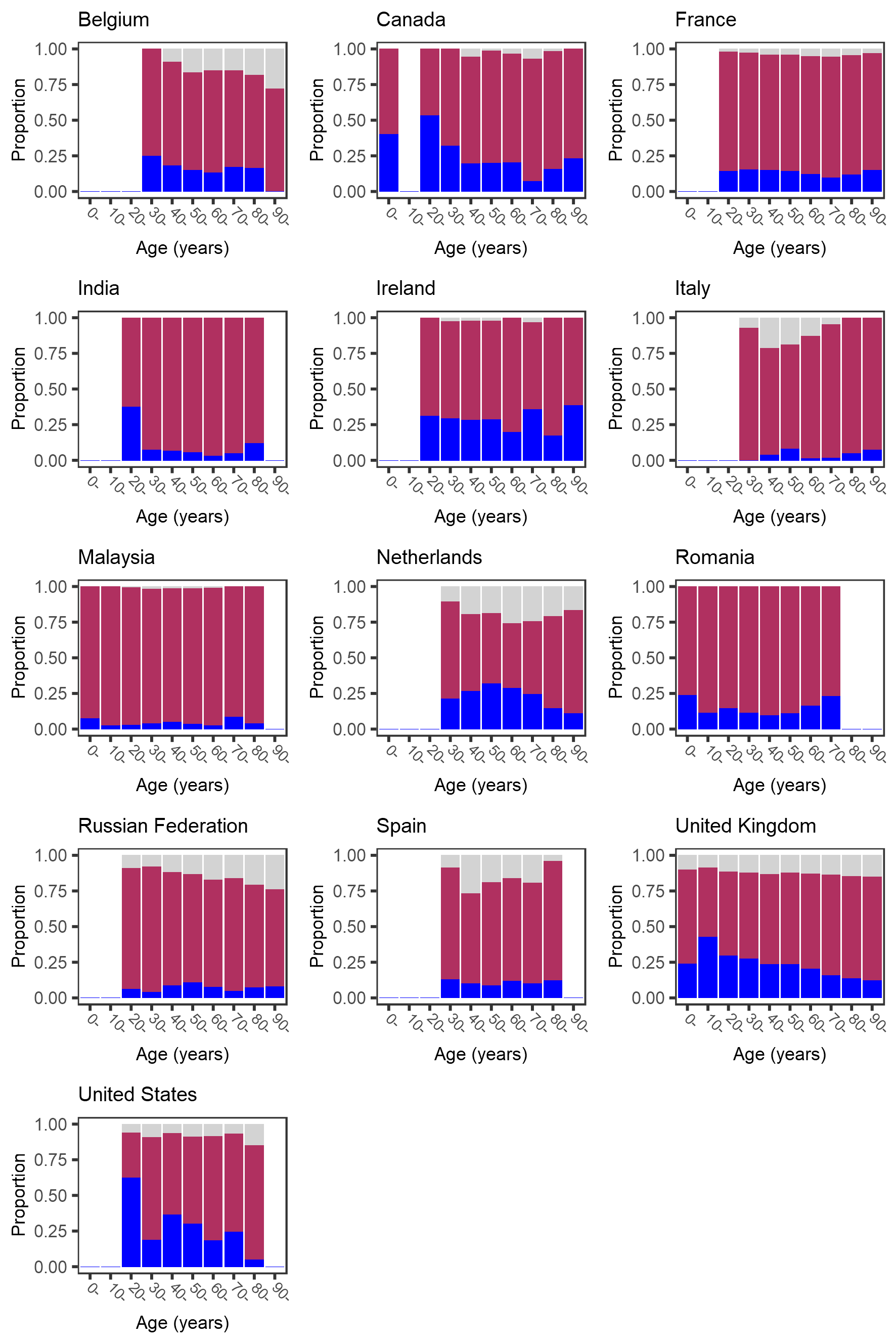

**Supplementary Figure 9. Age-specific prevalence of diarrhoea at hospital admission with covid‑19.**

Dark blue bars show symptom present, maroon bars show symptom absent, pale grey bars show missing data. Bars that would represent <10 individuals are omitted.

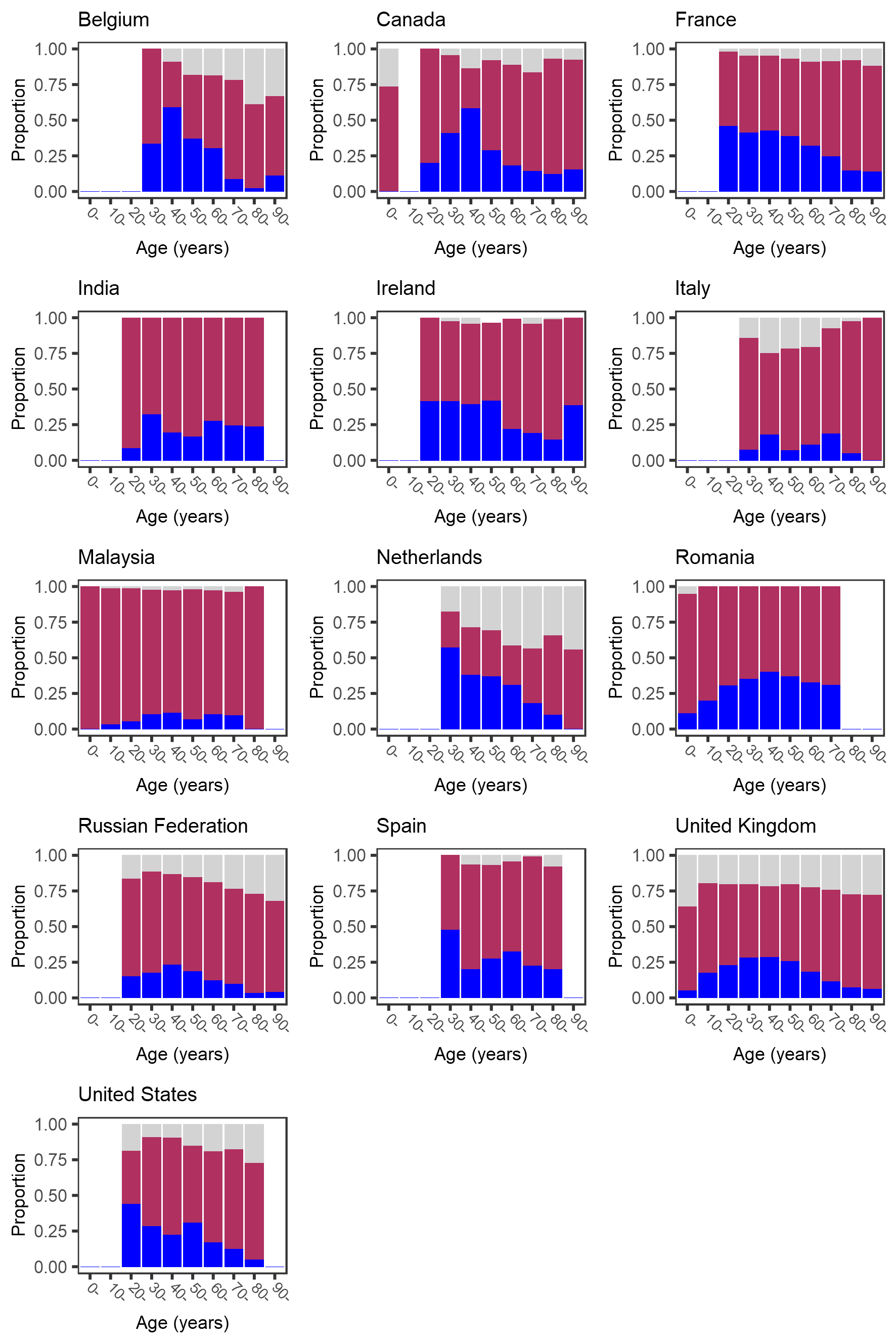

**Supplementary Figure 10. Age-specific prevalence of muscle pains at hospital admission with covid‑19.** Dark blue bars show symptom present, maroon bars show symptom absent, pale grey bars show missing data. Bars that would represent <10 individuals are omitted.

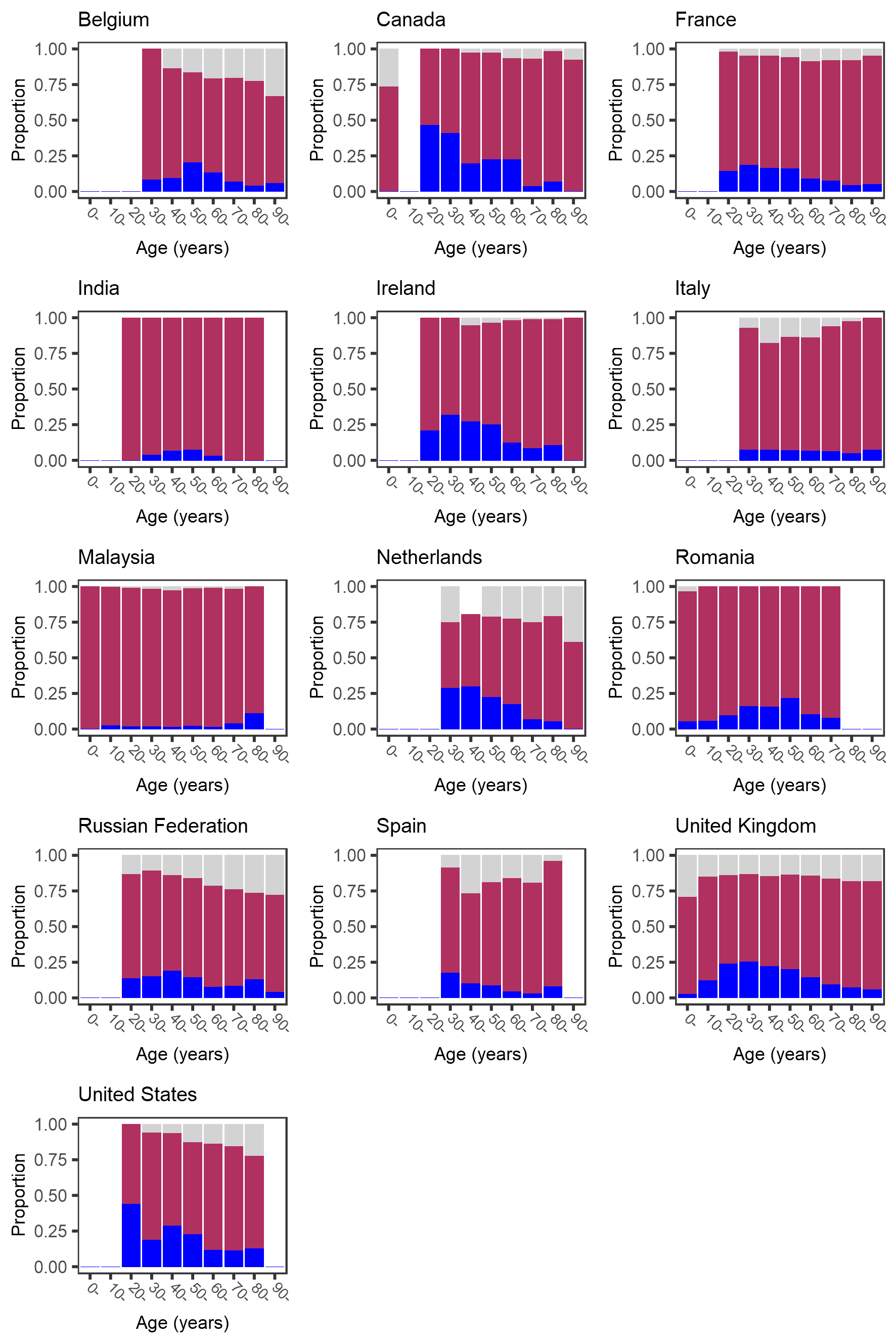

**Supplementary Figure 11. Age-specific prevalence of chest pain at hospital admission with covid‑19.** Dark blue bars show symptom present, maroon bars show symptom absent, pale grey bars show missing data. Bars that would represent <10 individuals are omitted.

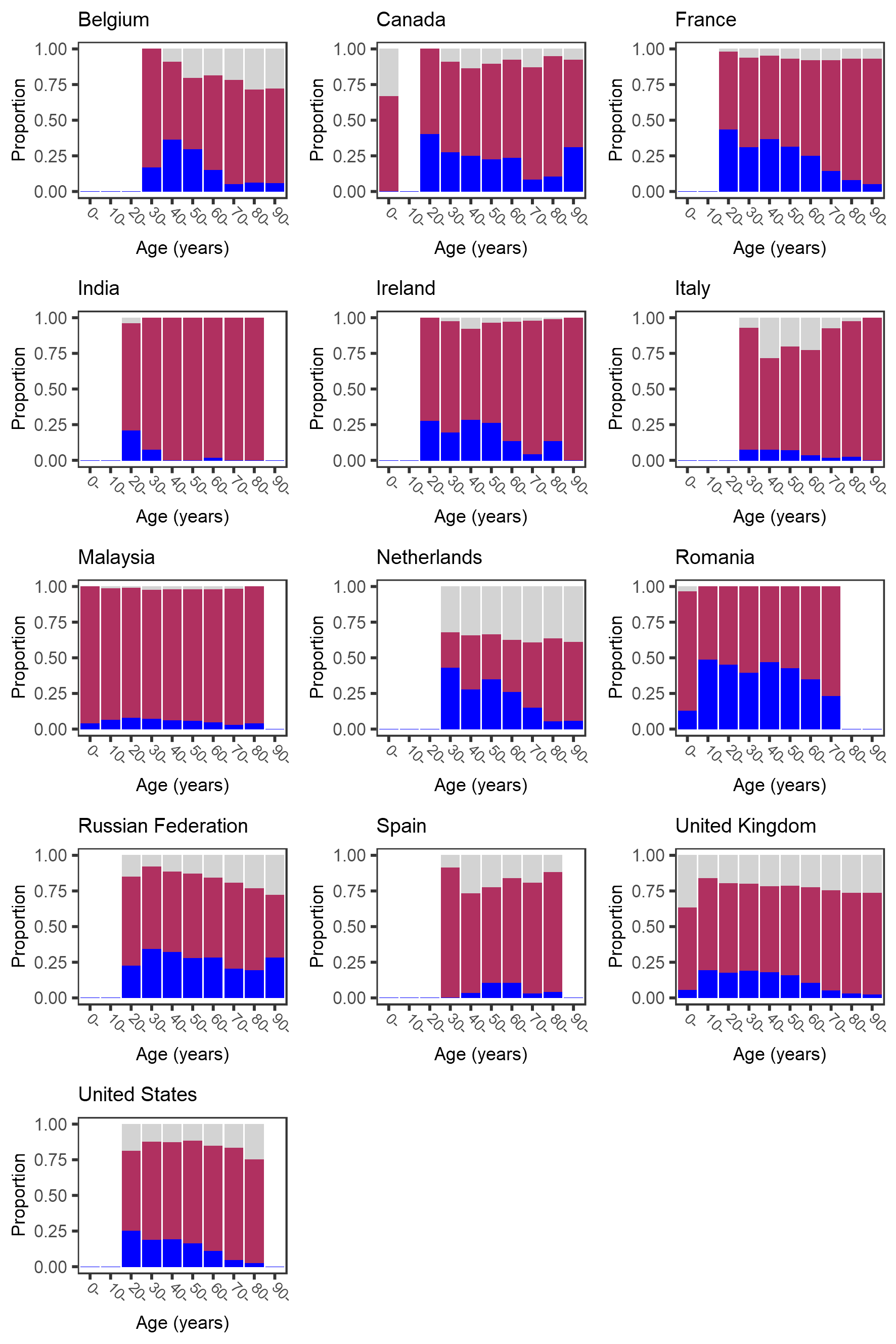

**Supplementary Figure 12. Age-specific prevalence of headache at hospital admission with covid‑19.** Dark blue bars show symptom present, maroon bars show symptom absent, pale grey bars show missing data. Bars that would represent <10 individuals are omitted.

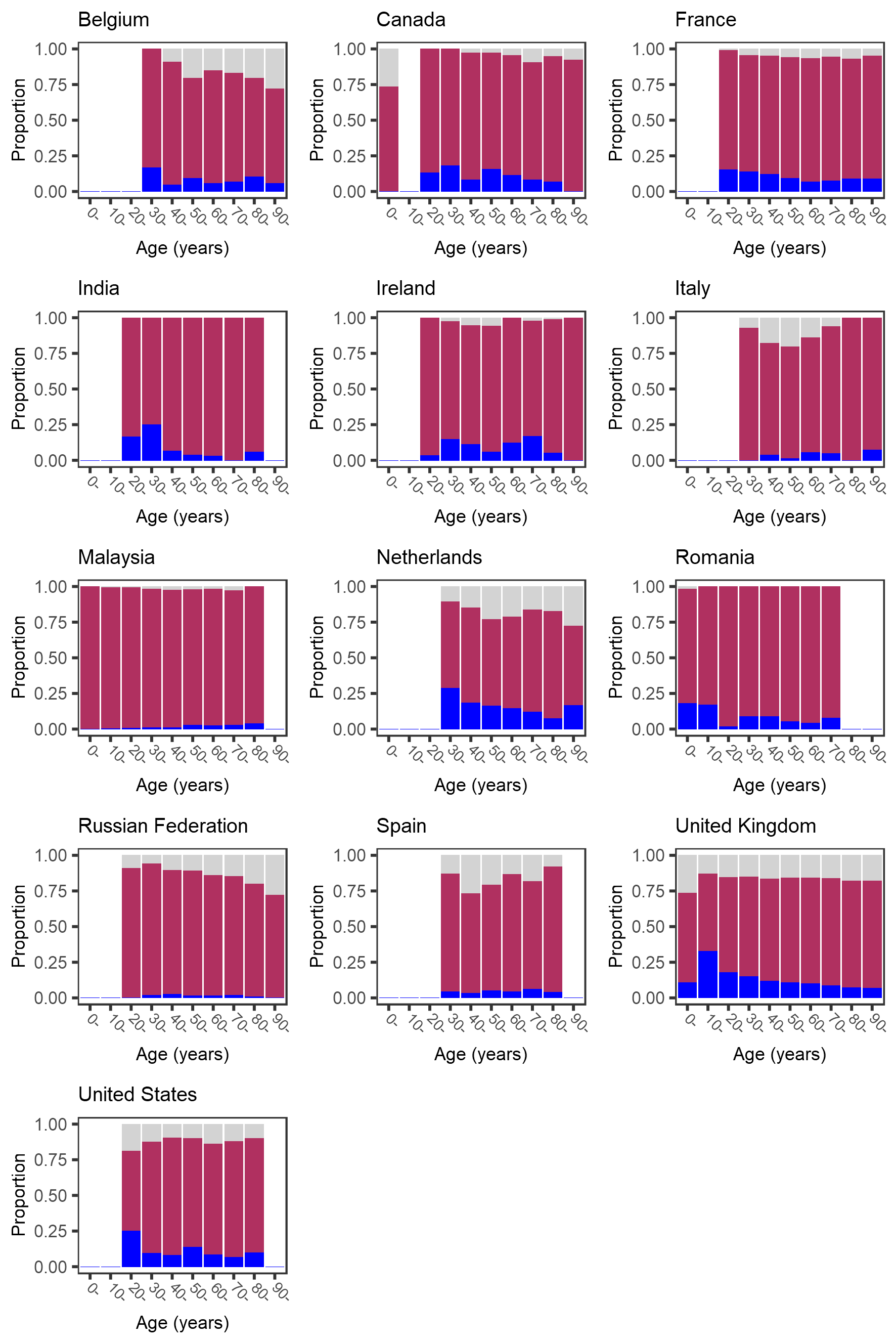

**Supplementary Figure 13. Age-specific prevalence of abdominal pain at hospital admission with covid‑19.** Dark blue bars show symptom present, maroon bars show symptom absent, pale grey bars show missing data. Bars that would represent <10 individuals are omitted.

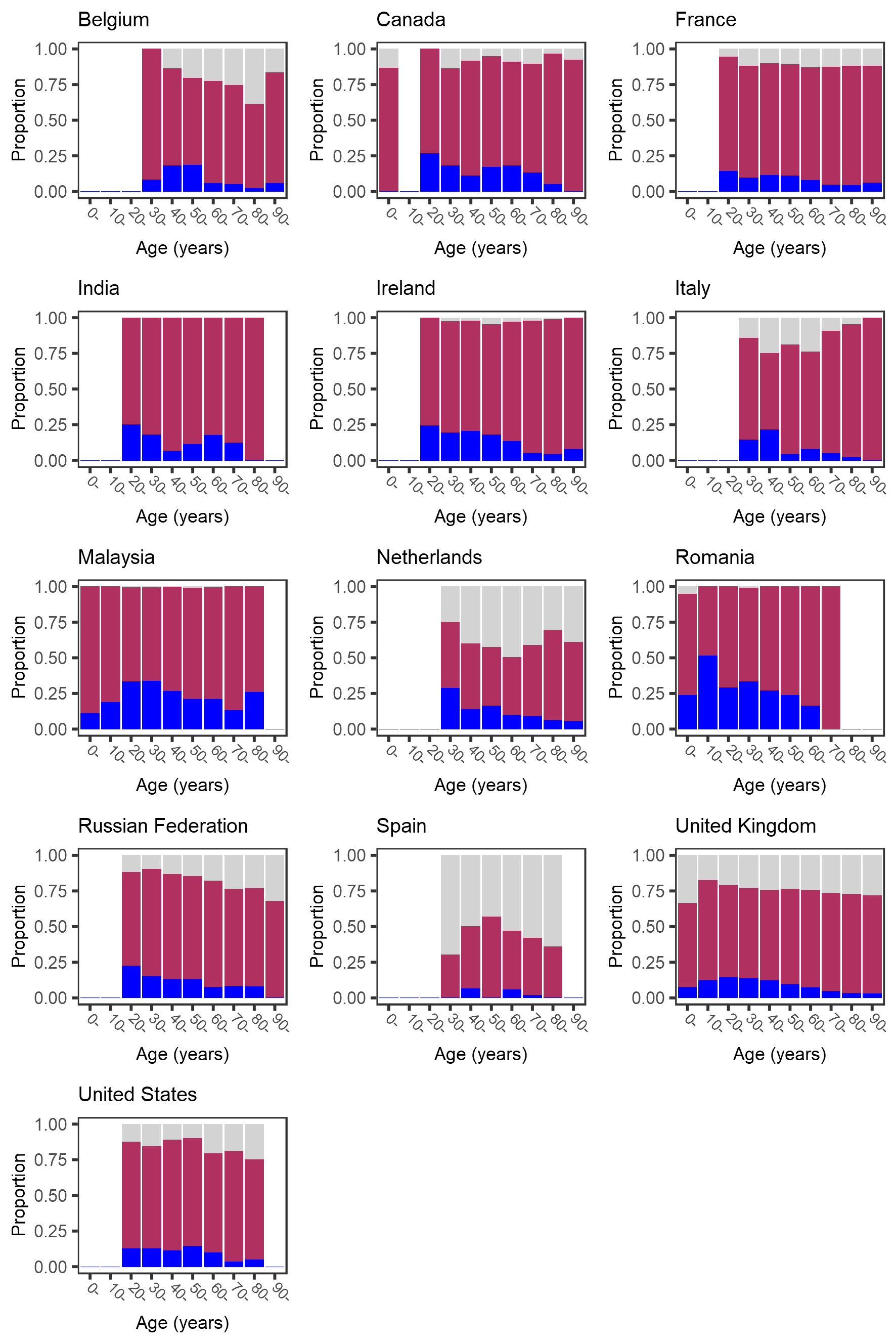

**Supplementary Figure 14. Age-specific prevalence of sore throat at hospital admission with covid‑19.**

Bars that would represent <10 individuals are omitted.

**CRediT author statement (based on Brand et al., 2015, doi: 10.1087/20150211)**

**Conceptualization**: (*study*) J. Kenneth Baillie, Gail Carson, Peter Horby, Laura Merson, Piero L. Olliaro, Malcolm G. Semple; (*this analysis*) Piero L. Olliaro, Mark G. Pritchard; **Methodology**: Emmanuelle A. Dankwa, Natalie Elkheir, Matthew Hall, Ewen M. Harrison, Antonia Ying Wai Ho, Christiana Kartsonaki, Kenneth A. McLean, Piero L. Olliaro, Mark G. Pritchard, Amanda Rojek, Louise Sigfrid; **Software and analysis**: Mark G. Pritchard; **Investigation**: Abdulrahman Al-Fares, Adrian Ceccato, Adrien Lemaignen, Adrienne Chan, Agamemnon Bakakos, Agnès Meybeck, Agnès Sommet, Agostinho Monteiro, Aine Mccarthy, Aisling Gavin, Al-Awwab Dabaliz, Albert Groenendijk, Alberto Cucino, Aldric Manuel, Alejandro Martin-Quiros, Aleksander Rygh Holten, Aleksandr Beljantsev, Alessandra Notari, Alexandra Bedossa, Alexandra Ducancelle, Alexandra Sperry, Ali Ait Hssain, Amanda Rojek, Amandine Gagneux-Brunon, Ami Stuart, Amna Faheem, Ana Catarino, Ana Freitas Ribeiro, Ana Hipólito-Reis, Ana Lúcia Rios, Ana Marques, Ana Martins, Ana Motos, Ana Pinho Oliveira Anca Streinu-Cercel, Anders Benjamin Kildal, André Cabie, André Dias, André Roberto, Andrea Dell’amore, Andrea Kelly, Andrea Rossanese, Andrea Villoldo, Andreas Lind, Andrés Orquera, Andrew Acker, Andrew Law, Andy Dong, Angel Sanchez-Miralles, Anika Atique, Anja Grosse Lordemann, Anjellica Chen, Anna Beltrame, Anna Greti Everding, Anne-Sophie Boureau, Anne-Sophie Resseguier, Anne Conrad, Anne Guillaumot, Anne Margarita Dyrhol-Riise, Ansley Hamer, Antoine Cheret, Antoine Kimmoun, Antoine Merckx, Anton Prinssen, Antonio Loforte, António Mesquita, Aquiles Henriquez-Trujillo, Arianne Joy Corpuz, Arie Zainul Fatoni, Arnaud Scherpereel, Asad Usman, Asfia Sultana, Asgar Rishu, Asma Moin, Audrey Barrelet, Bailey Cassandra, Barbara Wanjiru Citarella, Bénédicte Lefebvre, Benigno Ferreira, Benjamin Lefevre, Benjamin Smood, Benjamine Sarton, Benoît Gachet, Benoît Roze, Benoît Thill, Bernard Cholley, Bernardo Neves, Bernhard Rössler, Bertrand Dussol, Birgitte Stiksrud, Blake Mergler, Blandine Rammaert, Brenda Reeve, Brian Marsh, Brigitte Elharrar, Briseida Parra, Bruno Levy, Bryan Whelan, Budha Charan Singh Oinam, Caitriona Cody, Cameron J. Fairfield, Camille Bouisse, Camille Chassin, Carla Eira, Carlos M. Luna, Carlos Pimentel, Carmen Infante Dominguez, Carola Pierobon, Caroline Cullen, Caroline Kosgei, Caroline Martins Rego, Carrol Gamble, Catarina Silva, Catherine A. Shaw, Catherine Chakveatze, Catherine Chirouze, Catherine Marquis, Cécile Azoulay, Cécile Goujard, Cécile Mear-Passard, Cécile Tromeur, Cécile Yelnik, Cecilia Canepa, Cédric Joseph, Céline Michelanglei, César Vieira, Charbel Maroun Eid, Charles Crepy D’orleans, Charline Vauchy, Chih-Hsien Wang, Chloe Carpenter, Chloe Donohue, Chris Kandel, Christel Arnold-Day, Christian Buesaquillo, Christian Prebensen, Christian Rabaud, Christiana Kartsonaki, Christophe Fraser, Christophe Rapp, Claire Foley, Clara Flateau, Clara Mouton Perrot, Clare Jackson, Clark Owyang, Claudia Figueiredo-Mello, Claudia Milena Orozco-Chamorro, Claudia Montes, Claudina Cruz, Cleide Barrigoto, Clément Le Bihan, Clotilde Allavena, Colin Mccloskey, Cyril Le Bris, Daisuke Kasugai, Damien Bouhour, Daniel Glikman, Daniel Haber, Daniel Ivulich, Daniel Mathieu, Daniel Munblit, Daniel Perez, Daniel Plotkin, Daniela Guerreiro, Darshana Hewa Kandamby, David Molina, David Thomson, David Zucman, Davide Chiumello, Dawn Higgins, Deepali Kumar, Delphine Bergeaud, Delphine Lariviere, Demetrios Kutsogiannis, Denis Garot, Denis Malvy, Denise Richardson, Derek Murphy, Dewi Guellec, Diana Adrião, Diana Póvoas, Diego Fernando Bautista Rincon, Diego Franch-Llasat, Dimitra Melia Myrodia, Diogo Borges, Diogo Lopes, Domenico Luca Grieco, Dominique Deplanque, Dominique Luton, Dounia Bouhmani, Douwe F. Postma, Duong Bich Thuy, Edel Meaney, Edouard Soum, Eglantine Ferrand Devouge, Egle Saviciute, Eibhilin Higgins, Eka Yudha Lantang, Elena Gallego Curto, Elias Iosifidis, Elisa Demonchy, Elisabeth Adam, Elisabeth Botelho-Nevers, Elise Artaud-Macari, Ellen Shadowitz, Elodie Curlier, Elsa Nyamankolly, Else Quist-Paulsen, Ema Leal, Eman Al Qasim, Emanuele Durante Mangoni, Emily Somers, Emmanuel Roilides, Emmanuelle Dankwa, Emmanuelle Denis, Eric Gnall, Eric Oziol, Eric Senneville, Erlina Burhan, Erwan Fourn, Erwan L’her, Ethan Kurtzman, Eva Marwali, Eva Miranda Marwali, Eve Le Coustumier, Evert-Jan Wils, Ewa Talarek, Fabian Patauner, Fabrice Laine, Fanny Vuotto, Félix Djossou, Felwa Bin Humaid, Fernando Rainieri, Ferran Roche-Campo, Ferruccio Mele, Filipa Cardoso, Filipa Sequeira, Filipe Cardoso, Filomena Boccia, Fiona Griffiths, Firouzé Banisadr, Florence Jego, Florent Peelman, François-Xavier Catherine, François Bissuel, François Goehringer, François Lamontagne, François Martin Carrier, Frédérick D’aragon, Fredrik Müller, Gabriele Sales, Gabrielle Macheda, Gabrielle Ragazzo, Gary Leeming, Gennaro Martucci, Georges Le Falher, Geraldine Goco, Gezy Giwangkancana, Giles J Peek, Giorgia Montrucchio, Giovanna Occhipinti, Giovanna Panarello, Giulio Giovanni Cavalli, Gloria Crowl, Grégory Corvaisier, Gregory Purcell, Gretchen Lemmink, Gry Kloumann Bekken, Guillaume Martin-Blondel, Guillermo Giordano, Guylaine Castor-Alexandre, Gwenhaël Colin, Gyan Sandhu, Hajnal-Gabriela Illes, Hanna Jung, Hayato Taniguchi, Hayley Hardwick, Heidi Ammerlaan, Heidi Gruner, Hélène Salvator, Henry Castrillón, Hiba Zayyad, Hiroaki Hiraiwa, Hiroaki Shimizu, Hiroyuki Tanaka, Hodane Yonis, Hoi Ping Shum, Holger Neb, Hubert Tessier-Grenier, Hugo Inácio, Hugo Miranda Maldonado, Hugues Aumaitre, Hugues Cordel, Huynh Trung Trieu, Hwa Jin Cho, Ilka Engelmann, Imrana Khalid, Indrek Rätsep, Ioannis Trontzas, Ioannis Xynogalas, Ionna Deligiannis, Irfan Khan, Isabela Saba, Isabelle Delacroix, Isabelle Enderle, Isabelle Fabre, Ivo Castro, Jai Madhok, Jaime Hernandez-Montfort, James Lee, James Scott-Brown, Jan Cato Holter, Janet Harrison, Jarne Van Hattem, Jason Bouziotis, Jean-Benoît Arlet, Jean-Charles Gagnard, Jean-François Payen, Jean-Luc Diehl, Jean-Sébastien Hulot, Jean Baptiste Assie, Jeannet Bos, Jeff Powis, Jeffrey Aliudin, Jeffrey Javidfar, Jelmer Talsma, Jennifer Clarke, Jérémie Pasquier, Jérôme Dimet, Jess Gibson, Jimmy Ming-Yang Hsu, Jo Dalton, Joan Gómez-Junyent, Joana Ferrão, João Alves, João Camões, João Estevão, João Martins, João Oliveira, João Teixeira, Johann Auchabie, Jolanta Popielska, Jonathan Crump, Jonathan Golob, Jonathan Remppis, Jonathan Troost, Jordi Riera, Jorge Dantas, Jorge Fernandes, Jorge Paulos, Jorge Velazco, Jose Andres Calvache, José Casimiro, José Ernesto Vidal, Jose I Nunez, Jose M. Mandei, Jose Pedro Cidade, Joshua Solomon, Joy Ann Villanueva, Jp Connelly, Juan Fernado Masa Jimenez, Juan Jose Diaz Diaz, Julie Chas, Julien Jabot, Julien Moyet, Julien Poissy, Juliette Romaru, Junji Itai, Kai Zacharowski, Kalynn Kennon, Karen Delavigne, Karine Faure, Karine Risso, Karl Erik Müller, Karolina Krawczyk, Karolina Nowicka, Katharina Weil, Kazali Enagnon Alidjnou, Keith Wille, Kenneth A. Mclean, Kévin Alexandre, Kévin Bouiller, Kévin Didier, Koji Hoshino, Konrad Zawadka, Konstantinos Kyriakoulis, Konstantinos Syrigos, Konstanty Szuldrzynski, Kota Hoshino, Kristian Tonby, Lara Absil, Lars Heggelund, Laura Marsh, Laura Merson, Laura Van Gulik, Lauren Deconninck, Laurence Bouillet, Laurence Maulin, Laurène Azemar, Laurent Bitker, Laurent Guilleminault, Laurent Lefebvre, Laurent Plantier, Laurent Richier, Leanne Hays, Lenka Petroušová, Letizia Lucia Florio, Liadain Reid, Liem Luong, Lionel Piroth, Lisa Norman, Lorenzo Bertolino, Louis Gerbaud Morlaes, Lovkesh Arora, Luca Brazzi, Lucia Moro, Lúcia Proença, Luís Bento, Luis Felipe Reyes, Luís Patrão, Luís Val-Flores, Luisa Quesada, Lukas Arenz, Magdalena Surovcová, Maggie Mechlin, Maïder Pagadoy, Maire-Laure Casanova, Malte Kohns Vasconcelos, Manuel Etienne, Maram Zahran, Marc Lambert, Marcel Van Den Berge, Marcela Vieira Freire, Mare Pejkovska, Margarida Torres, Margaux Isnard, Maria Amaral, Maria Boylan, Maria Joao Silva, Maria Pokorska-Spiewak, Maria Sousa Uva, Maria Toki, Mariana Cascão, Mariano Esperatti, Marie-Christine Carret, Marie Connor, Marie Lachatre, Marie Lacoste, Marie Lagrange, Marie Langelot-Richard, Marie Piel-Julian, Marie Rafiq, Marielle Boyer-Besseyre, Marielle Buisson, Marília Fernandes, Marina Lanza, Mário Ferraz, Mario Palacios, Marion Le Maréchal, Marion Zabbe, Mark G Pritchard, Marlène Murris, Marlene Santos, Marta Leal Santos, Marta Sousa, Martin Martinot, Martine Remy, Mary Copland, Massimo Antonelli, Massimo Gagliardi, Mathieu Blot, Mathieu Lesouhaitier, Mathieu Mattei, Matthew Cummings, Matthew Griffee, Matthew Hall, Matthieu Revest, Mauro Panigada, Maxime Hentzien, Maximilian Malfertheiner, Medhi Mezidi, Mélanie Roriz, Mia Callahan, Michael Schwameis, Michael Sonntagbauer, Michaela Barnikel, Michela Leone, Michelle Girvan, Michelle Smyth, Mirjam Evers, Mohamed Fayed, Mohammed El Sanharawi, Mohammed Quraishi, Moïse Machado, Monserrat Solis, Morgane Snacken, Moshe Matan, Murray Wham, Musharaf Sadat, Mylène Maillet, Nadia Malik, Nadia Ouamara, Nadia Saidani, Natalie Mc Evoy, Nataly Farshait, Nathalie Allou, Nathalie De Castro, Nathalie Dournon, Nathalie Pansu, Niamh Feely, Nicholas Rizer, Nicholas Sedillot, Nick Daneman, Nicky Van Der Vekens, Nicolas Benech, Nicolas Brozzi, Nicolas Carlier, Nicolas Terzi, Nidyanara Castanheira, Nikita A Nekliudov, Nina Buchtele, Nisreen Shiban, Nora Fuentes, Nuno Germano, Odile Launay, Olavi Maasikas, Olguta Lungu, Olivier Bouchaud, Olivier Epaulard, Olivier Lesens, Olivier Robineau, Olivier Sanchez, Orna Ni Choileain, Pablo Blanco-Schweizer, Paola Rodari, Parthena Savvidou, Pascal Granier, Patrick Rispal, Paul Campbell, Paul Le Turnier, Paul Loubet, Paula Custodio, Pauline Caraux-Paz, Pauline Yeung Ng, Pavan Kumar Vecham, Pedro Faria, Pedro Povoa, Peter Kiiza, Peter Van Der Voort, Phil Gallagher, Phoebe Ampaw, Pierre-Marc Villeneuve, Pierre Delobel, Pierre Tattevin, Pleun Terpstra, Polina Bugaeva, Prasan Kumar Panda, Pratima Sharma, Quentin Lepiller, Rachael Ellis, Rachael Mcconnochie, Rachida Ouissa, Rafael Mahieu, Raphaël Borie, Raul Neto, Razi Alalqam, Rebecca Hamidfar, Rebecca Holt, Renata Barbalho, Renato Reis, Riinu Pius, Rita Alves, Rob Fowler, Roberta Cavalin, Roberto Andini, Robin Kobbe, Rodrigo Diaz, Romain Decours, Romain Guery, Roman Ullrich, Ross Hendry, Rostane Gaci, Roxane Courtois, Rui Pereira, Ruth Lyons, Ruth Noemí Jorge García, Ryuzo Abe, Saad Nseir, Sabelline Bouchez, Sabina Mason, Sabine Croonen, Sally Shrapnel, Samuel Mcelwee, Sanne Wesselius, Santi Rahayu Dewayanti, Saptadi Yuliarto, Sara Clohisey, Sara Ventura, Sarah Cormican, Sarah Isgett, Sarah Macdonald, Sarah Mcdonald, Sarah Redl, Scott Pharand, Sean Keating, Segolène Greffe, Sergio Poli, Séverine Ansart, Shaun Thompson, Sheila Cárcel, Sheryl Ann Abdukahil, Shingo Adachi, Shinichiro Ohshimo, Shirin Tabrizi, Shirley Sarfo-Mensah, Silvia Castañeda, Silvia Duranti, Simão Rodeia, Simon-Djamel Thiberville, Simon Bessis, Simone Carelli, Simone Piva, Simreen Kaur Johal, Smaragdi Kalomoiri, Sofia Cardoso, Sofía Contreras, Sofia Prapa, Sophie Halpin, Sophie Mahy, Stanislas Rebaudet, Stéphane Fry, Stéphane Jaureguiberry, Stéphane Sallaberry, Stephanie-Susanne Stecher, Stephanie Nonas, Stephanie Roberts, Stephen Knight, Steven Van Lelyveld, Su Hwan Lee, Subbarao Elapavaluru, Sue Smith, Sung Min Cho, Susana Cabral, Susana Fernandes, Susanne Dudman, Suzanne Bennett, Sybille Bevilcaqua, Sylvain Diamantis, Sylvie Le Gac, Sylvie Lion-Daolio, Synne Jenum, Takako Akimoto, Taku Tanaka, Tamara Seitz, Tânia Sequeira, Tarek Elshazly, Tatiana Fonseca, Tawnya Ogston, Thibault Chiarabini, Thomas Guimard, Thomas Maitre, Thomas Perpoint, Thomas Staudinger, Tiago Isidoro, Tiffany Trouillon, Timo Brandenburger, Tiphaine Goulenok, Tjard Schermer, Tom Drake, Toshiki Yokoyama, Triona Downer, Valentine Campana, Valérie Gaborieau, Valérie Garrait, Valérie Gissot, Vanina Meyssonnier, Vegard Skogen, Véronique Lemee, Vicente Souza-Dantas, Victoria Manda, Victoria Shaw, Vincent Dinot, Vincent Dubee, Vincent Le Moing, Vincent Peigne, Vincent Prestre, Vincent Thibault, Vladislav Mihnovitš, Walter Picard, William Dechert, William Greenhalf, Wilna Oosthuyzen, Wim Dieperink, Wing Yiu Ng, Xavier Sánchez Choez, Yael Dishon, Yih-Sharng Chen, Ymkje Stienstra, Yoan Lavie-Badie, Yohan N’guyen, Younes Ait Tamlihat, Younes Kerroumi, Yuri Kida, Yusing Gu; **Resources and funding**: Abdulrahman Al-Fares, Adrian Ceccato, Adrian Streinu-Cercel, Adrienne Chan, Alejandro Martin-Quiros, Ami Stuart, Anders Benjamin Kildal, Andrea Dell’amore, Andreas Lind, Andrey A Svistunov, Andy Dong, Angel Asensio, Angel Sanchez-Miralles, Anna Greti Everding, Anne Margarita Dyrhol-Riise, Antonio Loforte, Arabella Fahy, Asad Usman, Asgar Rishu, Bairbre Mcnicholas, Bharath Kumar Tirupakuzhi Vijayaraghavan, Bianca Boxma, Bryan Whelan, Budha Charan Singh Oinam, Carla Eira, Charles Crepy D’orleans, Chris Kandel, Clark Owyang, Claudia Figueiredo-Mello, Claudio Duarte Fonseca, Cornelius Rau, Daisuke Kasugai, Darshana Hewa Kandamby, David Molina, David S. Y. Ong, David Thomson, Davide Chiumello, Deepali Kumar, Demetrios Kutsogiannis, Denis Butnaru, Detlef Kindgen-Milles, Diana Póvoas, Diego De Mendoza, Douwe F. Postma, Eka Yudha Lantang, Elena Gallego Curto, Elias Iosifidis, Emanuele Durante Mangoni, Emily Martin, Emily Somers, Emmanuel Roilides, Eric Gnall, Eva Marwali, Eva Miranda Marwali, Ewa Talarek, Fabian Patauner, Ferruccio Mele, Filipa Sequeira, François Lamontagne, François Martin Carrier, Frédérick D’aragon, Fredrik Müller, Gabriele Sales, Gabrielle Ragazzo, Gezy Giwangkancana, Giorgia Montrucchio, Giulio Giovanni Cavalli, Hans Martin Bosse, Heidi Ammerlaan, Helen Tuite, Hiroyuki Tanaka, Indrek Rätsep, Irfan Khan, Jai Madhok, Jan Cato Holter, Jeff Powis, Jeffrey Javidfar, Jia Wei, John Marshall, Jolanta Popielska, Jonathan Remppis, Jose Andres Calvache, Jose Pedro Cidade, Juan Fernado Masa Jimenez, Juan Jose Diaz Diaz, Kai Zacharowski, Karolina Nowicka, Keith Wille, Konrad Zawadka, Konstantinos Syrigos, Kota Hoshino, Lars Heggelund, Le Van Tan, Letizia Lucia Florio, Luca Brazzi, Luís Patrão, Malte Kohns Vasconcelos, Maria Angelica Rivera Nuñez, Maria Donnelly, Maria Pokorska-Spiewak, Maria Toki, Massimo Antonelli, Massimo Gagliardi, Matthew Cummings, Matthew Griffee, Matthew Pellan Cheng, Mauro Panigada, Max O’donnell, Mia Callahan, Michael Collins, Mireia Cantero, Mohamed Fayed, Mohammed Quraishi, Nagarajan Ramakrishnan, Nataly Farshait, Navy Lolong, Niamh Feely, Nick Daneman, Nina Buchtele, Nisreen Shiban, Olguta Lungu, Parthena Savvidou, Paul Campbell, Pedro Povoa, Peter S Timashev, Peter Van Der Voort, Petr Glybochko, Prasan Kumar Panda, Roberto Andini, Robin Kobbe, Rosanna De Rosa, Rosario Maria Torres Santos-Olmo, Sabina Mason, Santi Rahayu Dewayanti, Sheila Cárcel, Simone Piva, Stephanie-Susanne Stecher, Stephanie Nonas, Steven Van Lelyveld, Subbarao Elapavaluru, Susanne Dudman, Tala Al-Dabbous, Timo Brandenburger, Tjard Schermer, Vicente Souza-Dantas, Victor Fomin, Wim Dieperink, Yaseen Arabi, Yih-Sharng Chen, Ymkje Stienstra, Yuri Kida, The Western Australian Covid-19 Research Response, The PETAL Network Investigators; **Data curation**: Sadie Kelly, Kalynn Kennon, James Lee, Laura Merson, Daniel Plotkin, Samantha Strudwick**; Writing - original draft**: Mark G. Pritchard, *with* Gail Carson, Ewen M. Harrison, Antonia Ying Wai Ho, Piero L. Olliaro, C. Russell, Louise Sigfrid; **Writing - review and editing**: All authors; **Visualization**: Mark G. Pritchard, **Supervision**: A.A. Roger Thompson, Abdulrahman Al-Fares, Adrian Ceccato, Adrian Streinu-Cercel, Adrienne Chan, Alberto Zanella, Alejandro Martin-Quiros, Aleksander Rygh Holten, Alessandra Notari, Alexander J. Mentzer, Alexander Zoufaly, Alexandra Coelho, Alexandre Gaymard, Alexandre Hoctin, Alexandros Cavayas, Ali Ait Hssain, Alison M. Meynert, Alistair Nichol, Alpha Diallo, Alphonsine Diouf, Ami Stuart, Amina Meziane, Ana Da Silva Filipe, Ana Marques, Ana Pinho Oliveira Anca Streinu-Cercel, Anders Benjamin Kildal, Andrea Dell’amore, Andreas Lind, Andrew Law, Angel Asensio, Angel Sanchez-Miralles, Anissa Chair, Anna Beltrame, Anne Margarita Dyrhol-Riise, Anne Mccarthy, Annelies Verbon, Annemarie B. Docherty, Antoine Khalil, Antonia Ying Wai Ho, Antonio Arcadipane, Antonio Loforte, Antonio Pesenti, Aquiles Henriquez-Trujillo, Arabella Fahy, Arthur Garan, Asad Usman, Asgar Rishu, Aurélie Papadopoulos, Aurélie Veislinger, Aurélie Wiedemann, Bairbre Mcnicholas, Beatrice Alex, Bénédicte Rossignol, Benjamin Bach, Benoit Visseaux, Bernhard Rössler, Bharath Kumar Tirupakuzhi Vijayaraghavan, Birgitte Stiksrud, Brenda Reeve, Brian Marsh, Bruno Lina, Bryan Whelan, Caitriona Cody, Cameron J. Fairfield, Camille Couffignal, Carine Roy, Carlos M. Luna, Caroline Semaille, Carolline De Araújo Mariz, Carrol Gamble, Catherine A. Shaw, Céline Dorival, Charbel Maroun Eid, Charlene Da Silveira, Charlotte Summers, Chloe Donohue, Christel Arnold-Day, Christelle Paul, Christelle Tual, Christl A. Donnelly, Christopher A. Green, Claire Andrejak, Claire Levy-Marchal, Clare Jackson, Clark D. Russell, Clark Owyang, Claudia Figueiredo-Mello, Colin McCloskey, Coralie Khan, Coralie Tardivon, Cornelius Rau, Daisuke Kasugai, Daniel Glikman, Daniel Munblit, Daniel Plotkin, David Dean, David L. Robertson, David S. Y. Ong, David Stuart, David Thomson, Davide Chiumello, Debby Bogaert, Deepali Kumar, Dehbia Benkerrou, Delphine Bachelet, Demetrios Kutsogiannis, Denis Malvy, Derek Murphy, Diana Póvoas, Diane Descamps, Diego De Mendoza, Diego Fernando Bautista Rincon, Dominique Deplanque, Dorothea Rosenberger, Douwe F. Postma, Eder Caceres, Edward Wilson Grandin, Egle Saviciute, Eka Yudha Lantang, Elena Gallego Curto, Elena Molinos, Elias Iosifidis, Ellen Shadowitz, Else Quist-Paulsen, Emanuele Durante Mangoni, Emily Martin, Emily Somers, Emma C. Thomson, Emmanuel Roilides, Eric D’ortenzio, Eric Gnall, Erlina Burhan, Esteban Garcia-Gallo, Ethan Kurtzman, Eva Marwali, Eva Miranda Marwali, Evelyne Kestelyn, Evert-Jan Wils, Ewen M. Harrison, Fernando Maltez, Filipa Sequeira, Filomena Boccia, Fiona Griffiths, Florentia Kaguelidou, France Mentré, François-Xavier Lescure, François Angoulvant, François Bompart, François Dubos, François Lamontagne, François Martin Carrier, François Téoulé, Frédérick D’aragon, Fredrik Müller, Gabriele Sales, Gabrielle Ragazzo, Gail Carson, Gary Leeming, Georgios Pollakis, Gerard Curley, Gezy Giwangkancana, Giacomo Grasselli, Giles J Peek, Gilles Peytavin, Giorgia Montrucchio, Giuseppe Foti, Graham S. Cooke, Gregory Purcell, Guillaume Lingas, Hanna Renk, Hans Martin Bosse, Helen Tuite, Hélène Esperou, Henk Vanoverschelde, Hervé Le Nagard, Hiba Zayyad, Hiroyuki Tanaka, Huda Alfoudri, Hugo Miranda Maldonado, Hugo Mouquet, Hwa Jin Cho, Ignacio Martin-Loeches, Indrek Rätsep, Ingrid G. Bustos, Ioana Grigoras, Irfan Khan, Isabelle Gorenne, Isabelle Hoffmann, Ithan D. Peltan, J. Kenneth Baillie, Jacobien Ellerbroek, Jade Ghosn, Jai Madhok, Jake Dunning, James Lee, James Scott-Brown, Jan-Erik Berdal, Jan Cato Holter, Jan Heerman, Janet Harrison, Janet T. Scott, Jarne Van Hattem, Javier Osatnik, Jean-Christophe Goffard, Jean-François Timsit, Jean Christophe Lucet, Jeannet Bos, Jeff Powis, Jeffrey Javidfar, Jérémie Guedj, Jimmy Mullaert, Jo Dalton, John G Laffey, John Marshall, Jolanta Popielska, Jonathan Golob, Jonathan Remppis, Jordi Riera, Jorge Velazco, Jose Andres Calvache, Jose M. Mandei, Jose Pedro Cidade, Juan Fernado Masa Jimenez, Juan Jose Diaz Diaz, Juan Pablo Horcajada, Julian A. Hiscox, Justine Pages, Kai Zacharowski, Keith Wille, Kenneth A. Mclean, Kevin Katz, Kollengode Ramanathan, Konstantinos Syrigos, Konstanty Szuldrzynski, Kota Hoshino, krishna Bhavsar, Kristian Tonby, Kusum Menon, Lance C.w. Turtle, Lars Heggelund, Lars Siegfrid Maier, Laura Marsh, Laura Merson, Laura Van Gulik, Laurent Abel, Lila Bouadma, Lina Morales-Cely, Lisa Norman, Lorenzo Bertolino, Louise Sigfrid, Lovkesh Arora, Luca Brazzi, Lucian Durham III, Luis Felipe Reyes, Lysa Tagherset, Mahdad Noursadeghi, Malcolm G Semple, Malte Kohns Vasconcelos, Manuel Etienne, Manuel Rosa-Calatrava, Marc Csete, Marcel Van Den Berge, Maria Donnelly, Maria Toki, Maria Zambon, Marie-Capucine Tellier, Marie-Pierre Debray, Marie Connor, Marina Esposito-Farese, Marina Mambert, Marine Beluze, Marion Noret, Marion Schneider, Mark Joseph, Marlice Van Dyk, Martina Hennessy, Massimo Antonelli, Massimo Palmarini, Mathilde Desvallée, Matthew Cummings, Matthew Griffee, Matthew Pellan Cheng, Maude Bouscambert, Mauro Panigada, Max O’Donnell, Maximilian Malfertheiner, Meera Chand, Mehul Desai, Menaldi Rasmin, Michael Collins, Michael Schwameis, Michelle Girvan, Minerva Cervantes-Gonzalez, Minh Le, Mireia Cantero, Mohamed Fayed, Mohammad Shamsah, Morgane Gilg, Moshe Matan, Murray Wham, Nadège Neant, Nadia Ettalhaoui, Nagarajan Ramakrishnan, Nassima Si Mohammed, Nathalie Gault, Nathan Peiffer-Smadja, Nguyen Van Vinh Chau, Niamh Feely, Nicholas Price, Nick Daneman, Nicolas Brozzi, Nina Buchtele, Nobuaki Shime, Noémie Mercier, Noémie Vanel, Olivier Picone, Olivier Terrier, Oriane Puéchal, Oscar Hoiting, Ouifiya Kafif, Patrick Biston, Patrick Rossignol, Paul Klenerman, Pauline Yeung Ng, Pedro Povoa, Peter Horby, Peter Kiiza, Peter Openshaw, Philippe Jouvet, Philippine Eloy, Piero Olliaro, Prasan Kumar Panda, Quentin Le Hingrat, Rachael Mcconnochie, Rafael León, Raul Neto, Ravi Kant, Ricard Ferrer-Roca, Richard S. Tedder, Riinu Pius, Roberto Andini, Roberto Roncon-Albuquerque Jr, Robin Kobbe, Rodrigo Diaz, Romain Basmaci, Roman Ullrich, Ross Hendry, Rui Pereira, Ruth Jimbo-Sotomayor, Ruth Lyons, Ryan S. Thwaites, Ryuzo Abe, Sabina Kali, Sabine Croonen, Sally Shrapnel, Salma Jaafoura, Samira Laribi, Samreen Ijaz, Sandrine Couffin-Cadiergues, Santi Rahayu Dewayanti, Saptadi Yuliarto, Sara Clohisey, Sarah Mcdonald, Sarah Tubiana, Saye Khoo, Sean Keating, Sheila Cárcel, Shinichiro Ohshimo, Shiranee Sriskandan, Shona C. Moore, Simone Piva, Siri Goepel, Soizic Le Mestre, Sophie Halpin, Sophie Yacoub, Stephanie-Susanne Stecher, Stephanie Nonas, Stephanie Roberts, Stephen Knight, Steven Van Lelyveld, Stijn Van De Velde, Subbarao Elapavaluru, Sung Min Cho, Susana Fernandes, Susanne Dudman, Suzanne Bennett, Sylvie Behilill, Sylvie Van Der Werf, Synne Jenum, Tae Song, Takako Akimoto, Takayuki Ogura, Théo Trioux, Thomas Staudinger, Thushan De Silva, Tjard Schermer, Tom Drake, Tom Fletcher, Tom Solomon, Tristan Gigante, Vanessa Sancho-Shimizu, Ventzislava Petrov-Sanchez, Vicente Souza-Dantas, Victoria Shaw, Vincent Enouf, Wai Ching Sin, Wei Shen Lim, Wendy S. Barclay, William A. Paxton, William Dechert, William Greenhalf, Wilna Oosthuyzen, Xavier Duval, Yaseen Arabi, Yazdan Yazdanpanah, Yih-Sharng Chen, Ymkje Stienstra, Yuri Kida, Yves Levy; **Project administration**: A. A. Roger Thompson, Abdulrahman Al-Fares, Adrian Ceccato, Adrian Streinu-Cercel, Adrienne Chan, Aine McCarthy, Albert Groenendijk, Alberto Cucino, Alberto Uribe, Alejandro Martin-Quiros, Aleksander Rygh Holten, Alessandra Notari, Alexander J. Mentzer, Alexandra Coelho, Alexandre Gaymard, Alexandre Hoctin, Ali Ait Hssain, Alison M. Meynert, Allison Mcgeer, Alpha Diallo, Alphonsine Diouf, Amanda Rojek, Ami Stuart, Amina Meziane, Amna Faheem, Ana Da Silva Filipe, Ana Freitas Ribeiro, Ana Lúcia Rios, Ana Marques, Ana Motos, Anca Streinu-Cercel, Anders Benjamin Kildal, Andrea Angheben, Andrea Dell’amore, Andreas Lind, Andrew Law, Andrey A Svistunov, Andy Dong, Angel Asensio, Angel Sanchez-Miralles, Anissa Chair, Anna Ciullo, Anne-Marie Guerguerian, Anne Margarita Dyrhol-Riise, Annemarie B. Docherty, Antoine Khalil, Antoni Torres, Antonia Ying Wai Ho, Antonio Loforte, Aquiles Henriquez-Trujillo, Arabella Fahy, Arie Zainul Fatoni, Arthur Garan, Asad Usman, Asgar Rishu, Astarini Hidayah, Aurélie Papadopoulos, Aurélie Veislinger, Aurélie Wiedemann, Bairbre Mcnicholas, Barbara Wanjiru Citarella, Beatrice Alex, Bénédicte Rossignol, Benigno Ferreira, Benjamin Bach, Benoit Visseaux, Bernardo Neves, Bernhard Rössler, Bharath Kumar Tirupakuzhi Vijayaraghavan, Bianca Boxma, Brenda Reeve, Brian Marsh, Bruno Lina, Bryan Whelan, Budha Charan Singh Oinam, Caitriona Cody, Cameron J. Fairfield, Camille Couffignal, Caren Friedrich, Carine Roy, Carlo Giaquinto, Carlos Alexandre Antunes De Brito, Carlos M. Luna, Carolien Van Netten, Caroline Semaille, Carrol Gamble, Cassidy Codan, Catherine A. Shaw, Catherine L. Hough, Catherine Marquis, Cédric Laouénan, Ceila Maria Sant`Ana Malaque, Celina Turchi Martelli, Céline Dorival, Charlene Da Silveira, Charles Crepy D’orleans, Charlotte Summers, Chih-Hsien Wang, Chloe Donohue, Christelle Paul, Christelle Tual, Christian Buesaquillo, Christian Prebensen, Christiana Kartsonaki, Christophe Fraser, Christopher A. Green, Claire Andrejak, Claire Levy-Marchal, Clare Jackson, Clark D. Russell, Clark Owyang, Claudia Figueiredo-Mello, Claudia Milena Orozco-Chamorro, Colin Mccloskey, Conar O’Neil, Coralie Khan, Coralie Tardivon, Cornelius Rau, Cynthia Braga, Daisuke Kasugai, Daniel Glikman, Daniel Haber, Daniel Ivulich, Daniel Munblit, Daniel Perez, Daniel Plotkin, Darshana Hewa Kandamby, David Bellemare, David Dean, David L. Robertson, David Maslove, David Richardson, David Stuart, David Thomson, Davide Chiumello, Debby Bogaert, Deepali Kumar, Dehbia Benkerrou, Delphine Bachelet, Demetrios Kutsogiannis, Denis Butnaru, Denise Richardson, Derek Murphy, Diana Adrião, Diana Póvoas, Diane Descamps, Diego Fernando Bautista Rincon, Diego Rolando Morocho Tutillo, Domenico Luca Grieco, Dominic So, Dominique Deplanque, Dori-Ann Martin, Dorothy Breen, Douwe F. Postma, Duong Bich Thuy, Eder Caceres, Edmund Manning, Edward Wilson Grandin, Egle Saviciute, Eka Yudha Lantang, Elena Gallego Curto, Elena Molinos, Elias Iosifidis, Elisabeth Adam, Ellen Shadowitz, Emanuele Durante Mangoni, Emily Neumann, Emily Somers, Emma C. Thomson, Emmanuel Roilides, Emmanuelle Dankwa, Eric D’ortenzio, Eric Gnall, Erica Bak, Erlina Burhan, Esteban Garcia-Gallo, Ethan Kurtzman, Eva Miranda Marwali, Evelyne Kestelyn, Evert-Jan Wils, Ewen M. Harrison, Fernando Maltez, Ferran Roche-Campo, Filipa Sequeira, Fiona Griffiths, Flávio Marino, Florentia Kaguelidou, France Mentré, François-Xavier Lescure, François Angoulvant, François Bompart, François Dubos, François Lamontagne, François Lellouche, François Martin Carrier, François Téoulé, Frédérick D’aragon, Fredrik Müller, Gabriele Sales, Gabrielle Ragazzo, Gail Carson, Gary Leeming, Gayle Carney, Gennaro Martucci, Georgios Pollakis, Geraldine Goco, Gezy Giwangkancana, Giacomo Grasselli, Giles J Peek, Gilles Peytavin, Giorgia Montrucchio, Gloria Crowl, Graham S. Cooke, Gregory Purcell, Guillaume Lingas, Hanna Jung, Hanna Renk, Hannah Visser, Hayato Taniguchi, Hayley Hardwick, Heidi Ammerlaan, Heidi Gruner, Helene Esperou, Hélène Esperou, Henk Vanoverschelde, Hervé Le Nagard, Hiba Zayyad, Hiroaki Shimizu, Hiroyuki Tanaka, Hoi Ping Shum, Holger Neb, Huda Alfoudri, Hugo Miranda Maldonado, Hugo Mouquet, Huynh Trung Trieu, Hwa Jin Cho, Ignacio Martin-Loeches, Indrek Rätsep, Ingrid G. Bustos, Ioana Grigoras, Ionna Deligiannis, Irene Aragao, Irfan Khan, Isabelle Gorenne, Isabelle Hoffmann, Ivo Castro, J. Kenneth Baillie, Jacobien Ellerbroek, Jade Ghosn, Jai Madhok, Jake Dunning, James Joshua Douglas, James Lee, James Scott-Brown, Jan-Erik Berdal, Jan Cato Holter, Jan Heerman, Janet Harrison, Janet Liang, Janet T. Scott, Jaques Sztajnbok, Jarne Van Hattem, Javier Osatnik, Jean-Charles Preiser, Jean-Christophe Goffard, Jean-François Timsit, Jean Christophe Lucet, Jeannet Bos, Jeffrey Javidfar, Jelmer Talsma, Jérémie Guedj, Jimmy Mullaert, Jo Dalton, Joan Gómez-Junyent, Johannes Gebauer, John Fraser, John G Laffey, John Marshall, Jolanta Popielska, Jonathan Remppis, Jonathan Troost, Jordi Riera, Jorge Velazco, Jose Andres Calvache, José Ernesto Vidal, Jose I Nunez, Jose M. Mandei, Jose Miguel Cisneros Herreros, Jose Pedro Cidade, Juan Fernado Masa Jimenez, Juan Jose Diaz Diaz, Juan Macias Sanchez, Judit Villar, Julia Rodriguez Abreu, Julian A. Hiscox, Julian Chica, Justine Pages, Kai Zacharowski, Kalynn Kennon, Karl Erik Müller, Kate Ainscough, Kate Calligy, Kathy Brickell, Katie O’hearn, Keibun Liu, Keiki Shimizu, Keith Wille, Kenneth A. Mclean, Kenneth Mclean, Koji Hoshino, Konstantinos Syrigos, Konstanty Szuldrzynski, Kota Hoshino, Krishna Bhavsar, Kristian Tonby, Kusum Menon, Lance C.w. Turtle, Lars Heggelund, Laura Marsh, Laura Merson, Laura Van Gulik, Laurent Abel, Le Van Tan, Leonardo Salazar, Letizia Lucia Florio, Lila Bouadma, Lisa Norman, Lorenzo Bertolino, Louise Sigfrid, Lovkesh Arora, Luca Brazzi, Luis Felipe Reyes, Luisa Quesada, Lysa Tagherset, M Azhari Taufik, Machteld Van Der Feltz, Magdalena Surovcová, Mahdad Noursadeghi, Makoto Uchiyama, Malcolm G Semple, Malte Kohns Vasconcelos, Manuel Etienne, Manuel Rosa-Calatrava, Marc Csete, Marco Giani, Margarite Grable, Maria Donnelly, Maria Joao Silva, Maria Toki, Maria Zambon, Mariano Esperatti, Marie-Capucine Tellier, Marie-Pierre Debray, Marie Connor, Marina Esposito-Farese, Marina Mambert, Marine Beluze, Marion Noret, Marion Schneider, Mark Downing, Mark Joseph, Mark G. Pritchard, Marlene Santos, Marlice Van Dyk, Martina Hennessy, Mary Mone, Masahiro Fukuda, Masaki Yamazaki, Massimo Antonelli, Massimo Gagliardi, Massimo Palmarini, Mathilde Desvallée, Matthew Cummings, Matthew Griffee, Matthew Hall, Matthew Pellan Cheng, Maude Bouscambert, Mauro Panigada, Max O’Donnell, Maximilian Malfertheiner, Meera Chand, Mehul Desai, Mia Callahan, Mical Paul, Michael Piagnerelli, Michael Rose, Michael Schwameis, Michael Sonntagbauer, Michelle E Kho, Michelle Girvan, Minerva Cervantes-Gonzalez, Minh Le, Mireia Cantero, Mohamed Fayed, Monserrat Solis, Morgane Gilg, Moshe Matan, Murray Wham, Myung Jin Song, Nadège Neant, Nadia Ettalhaoui, Nassima Si Mohammed, Natalie Mcevoy, Nataly Farshait, Nathalie Gault, Nathan Peiffer-Smadja, Nguyen Van Vinh Chau, Niamh Feely, Nicholas Price, Nick Daneman, Nicky Van Der Vekens, Nicolas Brozzi, Nikita A Nekliudov, Nina Buchtele, Nisreen Shiban, Nobuya Kitamura, Noémie Mercier, Noémie Vanel, Oana Sandulescu, Olavi Maasikas, Oleksa Rewa, Olivier Picone, Olivier Terrier, Oriane Puéchal, Oscar Hoiting, Ouifiya Kafif, Pablo Blanco-Schweizer, Pamela Combs, Paola Rodari, Patricia Fontela, Patrick Breen, Patrick Rossignol, Paul Klenerman, Paul Young, Pauline Yeung Ng, Pawel Twardowski, Pedro Povoa, Peter De Vries, Peter Horby, Peter Kiiza, Peter Openshaw, Peter S Timashev, Petr Glybochko, Philippe Jouvet, Philippine Eloy, Piero Olliaro, Pierre-Marc Villeneuve, Pieter Depuydt, Pleun Terpstra, Polina Bugaeva, Prasan Kumar Panda, Quentin Le Hingrat, Rachael Mcconnochie, Rafael Freitas De Oliveira França, Raul Neto, Rebekha Garcia, Renee Douma, Richard S. Tedder, Riinu Pius, Rima Song, Rob Fowler, Roberta Cavalin, Roberto Andini, Robin Kobbe, Rodrigo Diaz, Romain Basmaci, Roman Ullrich, Ross Hendry, Rui Pereira, Ruth Jimbo-Sotomayor, Ruth Lyons, Ruth Noemí Jorge García, Ryan S. Thwaites, Ryuzo Abe, Sabina Kali, Sabina Mason, Sabine Cornelis, Salma Jaafoura, Samira Laribi, Samreen Ijaz, Sandrine Couffin-Cadiergues, Sanne Wesselius, Saptadi Yuliarto, Sara Clohisey, Sara Taleb, Sarah Macdonald, Sarah Mcdonald, Sarah Moore, Sarah Tubiana, Saye Khoo, Sean Keating, Sérgio Gaião, Shaun Thompson, Sheeba Hakak, Sheila Cárcel, Sherry Mcdermott, Shingo Adachi, Shinichiro Ohshimo, Shiranee Sriskandan, Shirley Sarfo-Mensah, Shona C. Moore, Simone Piva, Siri Goepel, Soizic Le Mestre, Sophie Halpin, Sophie Yacoub, Steffi Ryckaert, Stephan Schroll, Stephanie-Susanne Stecher, Stephanie Nonas, Stephanie Roberts, Stephen Knight, Steven Van Lelyveld, Stijn Van De Velde, Su Hwan Lee, Subbarao Elapavaluru, Sue Smith, Susana Fernandes, Susanne Dudman, Sylvie Behilill, Sylvie Van Der Werf, Tae Song, Tak-Hyuk Oh, Takako Akimoto, Takayuki Ogura, Tamara Seitz, Tatiana Fonseca, Terese Hammond, Théo Treoux, Thomas Staudinger, Thushan De Silva, Tjard Schermer, Todd Lee, Tom Drake, Tom Fletcher, Tom Solomon, Toshiki Yokoyama, Tristan Gigante, Vanessa Sancho-Shimizu, Vegard Skogen, Ventzislava Petrov-Sanchez, Vicente Souza-Dantas, Victor Fomin, Victoria Shaw, Vincent Enouf, Volkan Korten, Wai Ching Sin, Wei Shen Lim, Wendy S. Barclay, William A. Paxton, William Dechert, William Greenhalf, Wilna Oosthuyzen, Wing Yiu Ng, Xavier Duval, Yaseen Arabi, Yazdan Yazdanpanah, Yih-Sharng Chen, Ymkje Stienstra, Yuri Kida, Yves Levy.
